# Support needs, support use and perceived helpfulness of support in a cohort of people bereaved during the COVID-19 pandemic: Insights from a longitudinal survey

**DOI:** 10.64898/2026.01.06.26343556

**Authors:** Silvia Goss, Kali Barawi, Eileen Sutton, Rebecca Oates, Kathy Seddon, Stephanie Sivell, Mirella Longo, Alison Penny, Lucy E Selman, Emily Harrop

## Abstract

**Background:** The negative impacts of the COVID-19 pandemic on bereavement experiences and grief outcomes are relatively well documented. However, less is known about the evolving support needs of people bereaved during this time, what support they used and, crucially, how this support helped (or hindered) their coping. Beyond the pandemic context, evidence of how bereavement support needs relate to the use and perceived helpfulness of different types of support is needed to inform bereavement service provision and policies.

**Methods:** A longitudinal survey of people bereaved (any cause of death) during the COVID-19 pandemic in the UK, with data collected at four time points: baseline (T1; n = 711), c. 8 (T2; n = 384), 13 (T3; n = 295), and 25 (T4; n = 185) months post-bereavement. Support needs and use of informal and formal support sources were captured quantitatively at all time points, with the perceived helpfulness of support captured as free-text data and analysed thematically. Future support preferences were obtained in the final survey round (T4). At T2-T4, participants completed the Traumatic Grief Inventory (TGI-SR) to assess for indications of Prolonged Grief Disorder (PGD). Quantitative data were analysed descriptively.

**Results:** In early bereavement, findings demonstrate high levels of support needs across multiple domains, with the highest needs at baseline relating to managing grief and feelings surrounding the loss (49.9-59.8% across 4 items), feelings of anxiety and depression (52.8%) and social isolation and loneliness (52.0%). Support needs decreased markedly over time but persisted for those with indicated PGD, of whom 44.2% at two-years post-bereavement (T4) needed help with coming to terms with how their loved one died and with expressing their feelings and feeling understood and 41.2% with loneliness and isolation. Participants primarily relied on support from family and friends, followed by online community support and one-to-one support (counselling), with support use decreasing over time. Those with indicated PGD engaged with all types of support more frequently across all time points, yet a third (35.3%) did not access any formal bereavement or mental health support during the first year following their bereavement. The qualitatively described benefits of different informal and formal support sources reflected and demonstrated their efficacy in meeting these support needs, though inadequacies in support were also highlighted, particularly from friends and family. Support preferences for future bereavements under non-pandemic circumstances most commonly included family and friends (96%), in-person one-to-one support (75%), self-help resources (63%) and GPs (61%).

**Conclusion:** Findings illustrate the multiple and varied emotional and social support needs of bereaved people, which for those with indicated PGD endured over time. While these needs can be effectively met by informal and formal support-types, dissatisfaction with support from friends and family, and under-utilisation of formal support services amongst high-risk groups, suggests significant unmet need and missed opportunities. This highlights the importance of strengthening the support available to people within their networks and communities and improving access to a wide variety of support options, according to people’s needs and preferences.

## 1. Background

Bereavement is a near-universal experience, yet the way an individual grieves and copes is profoundly personal and impacted by many factors (1,2). The support people receive after a bereavement is recognised as one of the most amenable and influential factors in determining how an individual copes with their loss (3–6). Support can be enhanced and tailored to individual needs and circumstances but this requires an understanding of support needs, how they may change over time and what support is experienced as helpful/unhelpful in meeting these needs.

Public health approaches to bereavement care recognise that needs and levels of vulnerability/resilience within bereaved populations differ, and recommend a corresponding tiered approach to support (7–10). It is estimated that c.60% of bereaved people will cope without formal intervention, supported by friends, family and their communities (8). The first tier of the public health model reflects the needs of this majority group and is focused on strengthening support from social networks and providing information on grief and available support (7,8). Research shows that bereaved individuals primarily rely on support from family and friends and suggests that they find this informal social support the most helpful (3,11–13). Emotional support from close others enables expressions of grief (4,12,14,15) and shows empathy and care (3,4,14,16,17). Practical help with unfamiliar practical and administrative tasks following a bereavement (15,18) as well as with everyday tasks is also valued (16). However, bereaved people also commonly experience waning support, insensitive comments, a lack of empathy and understanding, not being listened to and feeling unable to express feelings and needs (3,4,12,13,16,17,19,20), which undermines the quality of support from friends and family and makes bereavement an even more isolating experience (21).

The second tier of the public health model includes structured, reflective support, typically in the form of peer support groups or individual counselling, and is considered appropriate for the estimated 30% of people who will have moderate needs for support (7,8,22). Third-tier support, including specialist grief, mental health and psychological interventions, should be targeted at the small minority (c.10%) of people at high risk of prolonged grief disorder (PGD) (7,8). Bereavement services commonly offer different types of support which cut across these three tiers, such as drop-in events, telephone support, peer support groups, individual and group counselling and specialist counselling for those with more complex needs (22–24). During the pandemic, online chat forums and support groups, web-based and self-help resources and online/telephone provision of counselling support became much more commonly used, reflecting the service adaptations that needed to be made in response to infection-control measures (25–27).

Consensus-based research has defined two core outcomes (28) and related support functions (29) for these different types of bereavement support interventions and services. In relation to the first outcome –‘ability to cope with grief’ (28), there was agreement that services should focus on enabling knowledge and understanding about grief reactions, provide reassurance and help people to normalise, find-meaning in and accept their grief and loss experiences (29). These support functions align with the construct of ‘loss oriented’ coping within the Dual Process Model (DPM) which describes how people oscillate between dealing with the loss of the deceased person (loss orientation (LO)) and negotiating the practical and psychosocial changes to their lives that occur as a result of the bereavement (restoration-orientation (RO)) (30). For the second outcome –‘quality of life and mental wellbeing’ (28) – which more closely resembles a restoration orientation (30), services are seen to have a role in helping people to become more optimistic and experience improvements in their identity, functioning and social relationships, while also providing opportunities for connection, comfort and mutual support (29). Practical and financial support and advice are also recognised as an important support function (28,29), needed to help people manage the many administrative/practical tasks and challenges that they face following a death (18,31).

Evidence from two mixed-methods reviews of individual and group-based interventions (22,32) describes impacts which reflect many of these support outcomes and functions (28,29). These benefits include; enabling loss and grief resolution, improved feelings of control and hopefulness, and – in group settings – reduced isolation and feelings of connectedness. Empathetic relationships and the opportunity to confide in those outside of existing networks were also valued for both individual and group-based models (22). Constant access to a supportive community and opportunities for remembrance were additionally noted for online communities (32). Negative impacts were minimal but included upset due to insensitive comments from others via unmoderated online forums (32) and difficulties expressing feelings in these settings (22). Interpersonal factors, such as positive relationships between group members, clients and counsellors were, however, noted as important success factors, as was the need for therapists and volunteers to possess appropriate cultural and experiential knowledge of community grief processes and norms (22). However, while this evidence on the impacts of different types of bereavement support aligns well with the outcomes and support functions described (28,29), less is known about the extent to which these outcomes/impacts reflect empirically observed support-needs of bereaved people, what types of support are best placed to meet these needs, and whether there are areas of need not addressed by informal and formal sources bereavement support, or reflected in the core outcomes and functions described for bereavement services (28,29).

Conducted during the Covid-19 pandemic in the UK, this mixed-methods study collected longitudinal survey data on bereavement support needs (using a scale derived from the core outcomes research described (28)) and experiences of and preferences for different types of informal and formal support, up to two years post-bereavement. Analysis of this data provides new insights into what support people need in the changing aftermath of their bereavement, and how effectively available support meets these needs. With comparisons also drawn between participants with and without indicated PGD, we add to pandemic and non-pandemic evidence on long-term bereavement support needs amongst high and lower-risk populations, with important implications for policy and practice.

## 2. Methods

### 2.1. Study design and aim

We conducted a longitudinal national survey investigating the experiences of a UK-based cohort of people bereaved during the Covid-19 pandemic during the first and second year after their loss (20,33–37). Data were collected at four time points: baseline (T1) (on average 5 months post-bereavement), and approximately 8-, 13- and 25-months post-bereavement (T2, T3, T4, respectively). This paper focuses on the reported support needs, support use and perceived impact of support captured across the four time points, and on future support preferences obtained at T4.

The Checklist for Reporting Results of Internet E-Surveys (CHERRIES; (38)) was followed (see Supplementary File 1).

### 2.2. Survey development and contents: Baseline survey and follow-up surveys

The baseline survey (T1) was an open web-based survey (20) developed by the study team, which included a public representative (KS), and study advisory group (see Supplementary File 2). It was piloted and refined with input from 16 bereaved public representatives. Non-randomised open and closed questions covered end-of-life care and bereavement experiences, and perceived needs for, access to and experiences of formal and informal bereavement support. Sociodemographic information was collected on the person who died and bereaved participants. Subsequent survey rounds (T2, T3, T4; see Supplementary Files 3 and 4), also developed and refined with the study advisory group, repeated questions on support needs and support use, alongside validated grief, wellbeing and social support measures (see (20,33)).

### 2.3. Outcome measures

#### Support needs

Participants were asked to rate on a scale from 1 (no support needed) to 5 (high level of support needed) how much support or help they had needed over recent months in relation to 13 items in three domains: i) Practical support, ii) Managing grief and iii) Quality of life and mental wellbeing. The support need items were author-developed, informed by previous studies (22,28) and further examined with an explanatory factor analysis using our baseline data (35).

#### Support use

At baseline, participants were asked what types of support they had used to cope with their loss, with the multiple-choice response options including family and friends, GPs and six forms of bereavement and mental health support provided by services (see Table 4). Subsequent surveys (T2, T3) were personalised (see Supplementary File 3) to follow-up on the use of these six bereavement-specific support types (where relevant), establishing whether participants were still accessing previously reported support and capturing if they had used any *new* bereavement support services, groups or resources since their previous survey. Support use in the final survey (T4; see Supplementary File 4) was again captured via a multiple-choice question, including all support sources from the baseline survey (i.e. no personalisation), plus two additional support sources: general community groups and written/audio resources. Information on any ‘other’ support used was also captured in open questions in all surveys and coded and categorised into the tiered groupings below. Types of recently accessed support were first identified separately for each of the four time points. To capture the highest level of support used, we categorised accessed support according to the three tiers of the public health model of bereavement (7):

- Tier 1a: Friends and family only
- Tier 1 b: Informal and information-based support: GP, helpline/instant webchat, online community support, informal support group, other (e.g. websites, podcasts, self-help material).
- Tier 2/3: Formal bereavement services and mental health support: One-to-one support/counselling, bereavement support group/group counselling, specialist mental health support.

#### Future support preferences

In the final survey round (T4), participants were asked about the types of support they would prefer if they were to experience another close bereavement in the future in non-pandemic circumstances. Participants used 5-point rating scales ranging from ‘strongly like’ to ‘strongly dislike’ to indicate their preference/appreciation for different types of informal, community-based and formal support. Where appropriate, participants were asked to provide separate ratings for in-person vs. virtual (i.e. via video call) support options.

#### Prolonged Grief Disorder

For each of the above quantitative outcomes, we also report descriptive results comparing those with indicated Prolonged Grief Disorder (PGD) and those without, based on completion of the Traumatic Grief Inventory (self-report version; TGI-SR;(39,40)) at surveys T2-T4 (see (33) for further details on the use and analysis of the TGI-SR in this study).

#### Free-text data: Support needs and perceived helpfulness of support

After rating their support needs and indicating the types of support they had recently used, participants at all four time points were asked to describe in free-text comments the ways in which the support had helped them. They were also asked to provide further details on any support needs that they had recently experienced/selected.

### 2.4. Study procedure

The baseline survey was administered via the JISC online survey platform (41) and was open from 28th August 2020 to 5th January 2021 (20,34–36). It was disseminated to a convenience sample via social and mainstream media, voluntary sector associations and bereavement support organisations, including those working with ethnically diverse communities. Organisations helped disseminate the voluntary (non-incentivised) survey by sharing on social media, webpages, newsletters, online forums and via direct invitations to potential participants. Hard-copy postal surveys were available on request.

The personalised follow-up surveys at T2 and T3 used individual survey links emailed directly to participants who had given consent to be contacted with further surveys. Labelled with their participant study ID, these surveys were individualised to follow up on the types of informal and formal bereavement support people had reported at baseline and/or subsequent surveys, respectively, and were emailed approximately 8- and 13-months post-bereavement. Where baseline surveys were completed at least five months post-death (or the date of death was not given), the second survey was sent out two months after the first survey was received.

### 2.5. Participants

Inclusion criteria for the initial baseline survey (T1): aged 18+ with the ability to consent; bereaved of a family member or close friend (any cause of death) during the Covid-19 pandemic after the introduction of social-distancing requirements in the UK (16/03/2020); death occurred in the UK.

### 2.6. Data analysis

Descriptive statistics, frequency tables and bar/line charts were used to describe demographic and categorical response data related to support needs, support use and future support preferences. Analysis was conducted using SPSS software, version 29.0.

Free text responses were analysed using inductive thematic analysis (42), involving line-by-line coding in NVivo 1.7 and identification of descriptive and analytical themes. A preliminary coding framework was developed based on a sample of survey responses in the initial baseline survey (T1) which was then revised and applied in an iterative process moving between the data and the analytical concepts to develop codes and themes grounded in the data (EH, ES, SG, KB). This involved independent double coding of 10% of the data set, regular discussion and cross-checking within the study team and review of final themes by the wider qualitative team. The same coding framework was applied to data collected at T2, T3 and T4 by SG, EH and RO, with continued regular discussions of further refinements to ensure rigour and reliability. We initially analysed the data separately for the four time points, before combining the data for the purpose of this paper. Themes and sub-themes are mapped onto the three support need areas described above: ‘Practical support’, ‘Managing grief’ and ‘Quality of life and mental wellbeing’.

### 2.7. Ethical approval

The study was approved by Cardiff University School of Medicine Research Ethics Committee (SMREC 20/59) and conducted in accordance with the Declaration of Helsinki. All respondents provided informed consent.

## 3. Results

### 3.1. Sample characteristics

Tables 1 and 2 show the characteristics of the participants and the person who had died, respectively, for each of the four survey rounds. A total of 711 participants completed the initial baseline survey (also see (20)), on average 137 days after the date of death (median = 152 days or 5 months; range 1 to 279 days). Participants ranged in age from 18 to 90 years (M=50, SD=13). Most were female (n=628, 89%) and had experienced the death of either a parent (n=395, 56%) or a partner (n=152; 21%). Thirty-three participants self-identified as being from an ethnically diverse background (4.7%). The most frequently reported cause of death was a confirmed or suspected Covid-19 infection (n=311, 44%) or cancer (n=156; 22%), with 78% of participants (n=552) indicating that they felt the death had been unexpected.

**Table 1:** Characteristics of the bereaved person and PGD status (based on TGI-SR score).

|  | **T1**<br>**(n=711)** |  | **T2**<br>**(n=384)** |  | **T3**<br>**(n=295)** |  | **T4**<br>**(n=185)** |  |
| --- | --- | --- | --- | --- | --- | --- | --- | --- |
| **Age** | **Mean±SD [Mdn]**<br>50 ± 13 [50]<br>**Min-Max**<br>18-90<br>**Missing data**<br>8 |  | **Mean±SD [Mdn]**<br>51 ± 13 [52]<br>**Min-Max**<br>21-90<br>**Missing data**<br>3 |  | **Mean±SD [Mdn]**<br>51 ± 13 [52]<br>**Min-Max**<br>22-86<br>**Missing data**<br>4 |  | **Mean±SD [Mdn]**<br>53 ± 12 [55]<br>**Min-Max**<br>22-84<br>**Missing data**<br>3 |  |
| **TGI-SR score** |  |  | **Mean ± SD [Median]**<br>51.5 ± 14.6 [52]<br>**Min-Max**<br>22-90<br>**Missing data**<br>2 |  | **Mean ± SD [Median]**<br>48.5 ± 14.7 [48]<br>**Min-Max**<br>18-89<br>**Missing data**<br>0 |  | **Mean ± SD [Median]**<br>44.7 ± 15.1 [43]<br>**Min-Max**<br>20-90<br>**Missing data**<br>0 |  |
| **PGD status**<br>(TGI score ≥ 54) |  |  | **n** | **%** | **n** | **%** | **n** | **%** |
| Yes |  |  | 167 | 43.7% | 102 | 34.6% | 53 | 28.6% |
| No |  |  | 215 | 56.3% | 193 | 65.4% | 132 | 71.4% |
| Didn't respond/Missing data |  |  | 2 |  | 0 |  | 0 |  |
| **Gender Identity** | **n** | **%** | **n** | **%** | **n** | **%** | **n** | **%** |
| Man | 74 | 10.0% | 45 | 11.7% | 32 | 11.0% | 25 | 14.0% |
| Woman | 628 | 89.0% | 337 | 88.0% | 261 | 89.0% | 159 | 86.0% |
| Non-binary/Other | 5 | 0.7% | 1 | 0.3% | 1 | 0.3% | 0 | 0.0% |
| Didn't respond/Missing data | 2 |  | 1 |  | 1 |  | 1 |  |
| **Ethnicity** | **n** | **%** | **n** | **%** | **n** | **%** | **n** | **%** |
| White (total) | 676 | 95.3% | 369 | 96.3% | 287 | 97.6% | 180 | 97.8% |
| *White British* | 438 | 64.8% | 242 | 65.6% | 179 | 62.4% | 114 | 63.3% |
| *White English* | 111 | 16.4% | 68 | 18.4% | 51 | 17.8% | 31 | 17.2% |
| *White Welsh* | 41 | 6.1% | 21 | 5.7% | 17 | 5.9% | 13 | 7.2% |
| *White Scottish* | 40 | 5.9% | 19 | 5.1% | 20 | 7.0% | 10 | 5.6% |
| *White Northern Irish* | 22 | 3.3% | 10 | 2.7% | 9 | 3.1% | 5 | 2.8% |
| *White Irish* | 7 | 1.0% | 2 | 0.5% | 3 | 1.0% | 3 | 1.7% |
| *Any other white background* | 17 | 2.5% | 7 | 1.9% | 8 | 2.8% | 4 | 2.2% |
| Ethnically diverse (total) | 33 | 4.7% | 14 | 3.7% | 7 | 2.4% | 4 | 2.2% |
| *White and Black Caribbean* | 11 | 33.3% | 3 | 21.4% | 1 | 14.3% | 1 | 25.0% |
| *Black Caribbean* | 4 | 12.1% | 2 | 14.3% | 1 | 14.3% | 0 | 0.0% |
| *White and Black African* | 2 | 6.1% | 0 | 0.0% | 0 | 0.0% | 0 | 0.0% |
| *Arab* | 1 | 3.0% | 1 | 7.1% | 1 | 14.3% | 0 | 0.0% |
| *Bangladeshi* | 2 | 6.1% | 1 | 7.1% | 1 | 14.3% | 1 | 25.0% |
| *Pakistani* | 1 | 3.0% | 0 | 0.0% | 0 | 0.0% | 0 | 0.0% |
| *Indian* | 4 | 12.1% | 3 | 21.4% | 0 | 0.0% | 0 | 0.0% |
| *White and Asian* | 4 | 12.1% | 3 | 21.4% | 2 | 28.6% | 2 | 50.0% |
| *Any other Asian background* | 1 | 3.0% | 0 | 0.0% | 0 | 0.0% | 0 | 0.0% |
| *Any other mixed background* | 3 | 9.1% | 1 | 7.1% | 1 | 14.3% | 0 | 0.0% |
| Didn't respond/Missing data | 2 |  | 1 |  | 1 |  | 1 |  |
| **Religious Beliefs** | **n** | **%** | **n** | **%** | **n** | **%** | **n** | **%** |
| Buddhism | 8 | 1.2% | 4 | 1.1% | 2 | 0.7% | 1 | 0.6% |
| Christianity | 244 | 37.7% | 144 | 40.0% | 111 | 39.6% | 76 | 43.4% |
| Hinduism | 3 | 0.5% | 1 | 0.3% | 0 | 0.0% | 0 | 0.0% |
| Islamism | 5 | 0.8% | 2 | 0.6% | 2 | 0.7% | 2 | 1.1% |
| Judaism | 6 | 0.9% | 2 | 0.6% | 3 | 1.1% | 2 | 1.1% |
| Sikhism | 2 | 0.3% | 1 | 0.3% | 0 | 0.0% | 0 | 0.0% |
| Other | 5 | 0.8% | 3 | 0.8% | 3 | 1.1% | 2 | 1.1% |
| Spiritual but not religious | 68 | 10.5% | 43 | 11.9% | 31 | 11.1% | 18 | 10.3% |
| Agnostic | 25 | 3.9% | 9 | 2.5% | 8 | 2.9% | 4 | 2.3% |
| Spiritual or religious (total) | 349 | 53.9% | 203 | 56.4% | 155 | 55.4% | 103 | 58.9% |
| No religious beliefs | 298 | 46.1% | 151 | 41.9% | 125 | 44.6% | 72 | 41.1% |
| Didn't respond/Missing data | 64 |  | 24 |  | 15 |  | 10 |  |
| **Highest Qualification** | **n** | **%** | **n** | **%** | **n** | **%** | **n** | **%** |
| None / GCSEs* | 108 | 15.0% | 50 | 13.0% | 35 | 12.0% | 25 | 14.0% |
| A-level / apprenticeship / ONC** | 132 | 19.0% | 68 | 17.7% | 48 | 16.0% | 24 | 13.0% |
| HND / University Degree*** | 468 | 66.0% | 266 | 69.3% | 212 | 72.0% | 136 | 74.0% |
| Didn't respond/Missing data | 3 |  | 0 |  | 0 |  | 0 |  |
| **Region** | **n** | **%** | **n** | **%** | **n** | **%** | **n** | **%** |
| England | 517 | 72.4% | 291 | 75.9% | 216 | 73.7% | 132 | 71.4% |
| Northern Ireland | 26 | 3.7% | 12 | 3.1% | 10 | 3.4% | 5 | 2.7% |
| Scotland | 53 | 7.5% | 28 | 7.3% | 28 | 9.5% | 17 | 9.2% |
| Wales | 63 | 8.9% | 37 | 9.7% | 26 | 8.8% | 20 | 10.8% |
| Didn't respond/Missing data | 52 |  | 16 |  | 15 |  | 11 |  |

All variables were captured at baseline only, except for the TGI-SR scores.
\*GCSE = General Certificate of Secondary Education for 15 and 16 year olds in the UK
\*\*A-levels = Advanced Level subject-based qualification for students in the UK aged 16 and above; ONC = Ordinary National Certificate (equivalent to A Levels)
\*\*\*HND = Higher National Diploma (vocational qualification provided by higher or further education colleges in the UK)

**Table 2:**
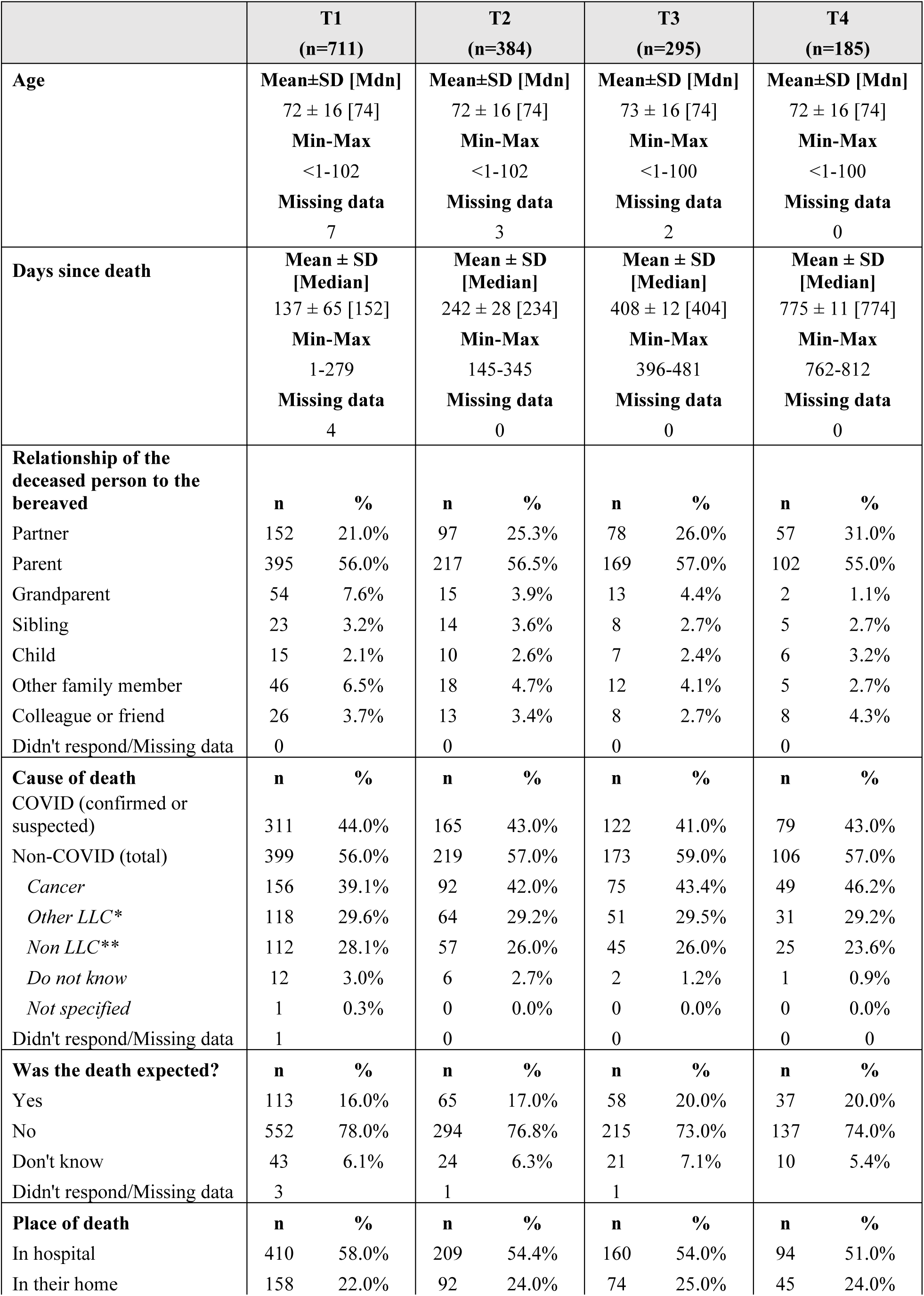
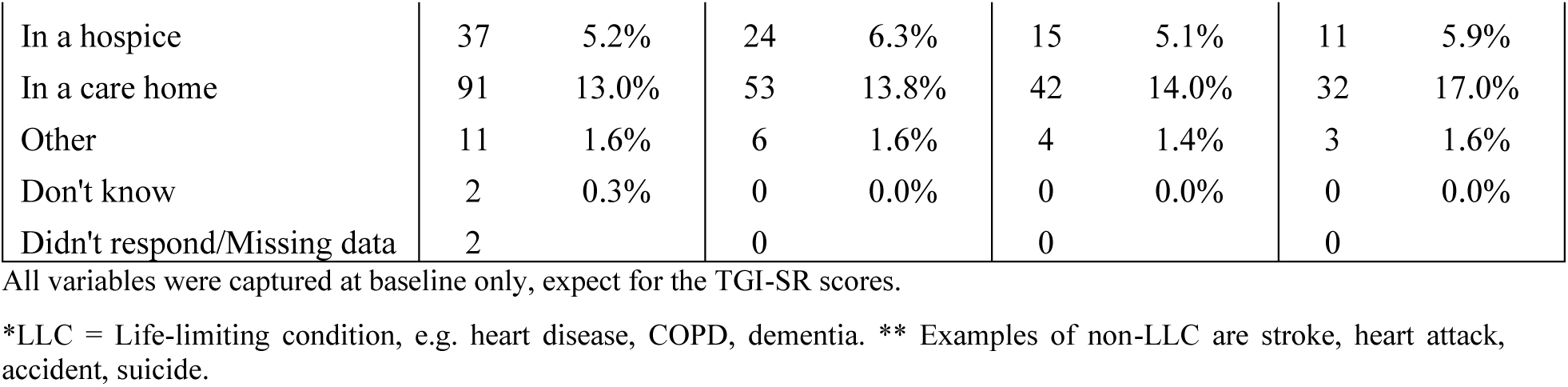
Characteristics of the person who had died.

The follow-up surveys were completed at T2 (n=384), T3 (n=295) and T4 (n=185); respectively, with 23.2% (n=165) of baseline respondents completing all three follow-up surveys. All second-round surveys were completed between 20/11/20 and 24/08/2021 and on average 242 days (median = 234 days or 8 months) after the date of death (range 145 to 345 days). All third-round surveys were completed between 04/05/2021 and 09/01/22 and on average 408 days (median = 404 or 13 months) after the date of death (range 396 to 481 days). All fourth-round surveys were completed between 17/05/2022 and 12/01/2023, on average 775 days (median = 774 or 25 months) after the date of death (range 762 days to 812 days) (see (33)). Overall, the sample composition remained relatively stable throughout the study. However, there was a tendency for the youngest and oldest participants to stop responding to the surveys, as well as those from ethnically diverse backgrounds and those with lowest qualification levels (see Table 1).

Mean TGI-SR scores decreased across the survey time points, from 51.5 at T2 to 48.5 at T3 and 44.7 at T4. At T2, 43.7% (n=167) met the threshold for indicated PGD (≥54;(39)), dropping to 34.6% at T3 (n=102) and 28.6% at T4 (n=53).

### 3.2. Support needs

As previously reported (20), fairly high or high levels of support need were commonly reported at baseline (T1). Within the ‘Managing grief’ domain, more than half of participants indicated needing support with ‘Dealing with my feelings about the way my loved one died’(59.8%), ‘Expressing my feelings and feeling understood’ (53.0%) and ‘Feeling comforted and reassured’ (51.7%), while within the ‘Quality of life/Mental wellbeing’ domain, the most common high-level support needs related to ‘Feeling of anxiety and depression’ (52.8%), ‘Loneliness and social isolation’ (52.0%) and ‘Regaining sense of purpose and meaning in life’ (46.7%). While the three most common support needs in these two domains remained the same at all three subsequent time points (T2/T3/T4), support needs gradually reduced over time and had dropped considerably by two-years post-bereavement (see Table 3).

**Table 3:** Levels of emotional and practical support needs across all four time points (T1 = baseline, 5-months post-bereavement (median); T2-T4 = 8-, 13- and 25-months post-bereavement, respectively).

|  |  | **T1**<br>**(n=711)** |  | **T2**<br>**(n=384)** |  | **T3**<br>**(n=295)** |  | **T4**<br>**(n=185)** |  |
| --- | --- | --- | --- | --- | --- | --- | --- | --- | --- |
|  |  | n | % | n | % | n | % | n | % |
| **Managing grief** |  |  |  |  |  |  |  |  |  |
| Dealing with my feelings about the way my loved one died | No or little support needed | 130 | 18.7% | 130 | 34.0% | 141 | 48.1% | 106 | 57.6% |
|  | Moderate level of support needed | 150 | 21.5% | 97 | 25.4% | 71 | 24.2% | 44 | 23.9% |
|  | Fairly high or high level of support needed | 417 | 59.8% | 155 | 40.6% | 81 | 27.6% | 34 | 18.5% |
|  | *Missing* | 14 |  | 2 |  | 2 |  | 1 |  |
| Expressing my feelings and feeling understood by others | No or little support needed | 160 | 23.1% | 133 | 35.1% | 139 | 47.6% | 108 | 59.0% |
|  | Moderate level of support needed | 166 | 23.9% | 108 | 28.5% | 89 | 30.5% | 46 | 25.1% |
|  | Fairly high or high level of support needed | 368 | 53.0% | 138 | 36.4% | 64 | 21.9% | 29 | 15.8% |
|  | *Missing* | 17 |  | 5 |  | 3 |  | 2 |  |
| Feeling comforted and reassured | No or little support needed | 148 | 21.6% | 101 | 26.5% | 118 | 40.7% | 107 | 58.5% |
|  | Moderate level of support needed | 183 | 26.7% | 127 | 33.3% | 99 | 34.1% | 48 | 26.2% |
|  | Fairly high or high level of support needed | 355 | 51.7% | 153 | 40.2% | 73 | 25.2% | 28 | 15.3% |
|  | *Missing* | 25 |  | 3 |  | 5 |  | 2 |  |
| Dealing with my feelings about being without my loved one | No or little support needed | 143 | 20.8% | 146 | 38.2% | 154 | 52.6% | 120 | 65.6% |
|  | Moderate level of support needed | 201 | 29.3% | 114 | 29.8% | 84 | 28.7% | 39 | 21.3% |
|  | Fairly high or high level of support needed | 342 | 49.9% | 122 | 31.9% | 55 | 18.8% | 24 | 13.1% |
|  | *Missing* | 25 |  | 2 |  | 2 |  | 2 |  |
| **Quality of life and mental wellbeing** |  |  |  |  |  |  |  |  |  |
| Feelings of anxiety and depression | No or little support needed | 180 | 26.0% | 162 | 42.3% | 164 | 56.2% | 108 | 58.7% |
|  | Moderate level of support needed | 146 | 21.1% | 89 | 23.2% | 52 | 17.8% | 43 | 23.4% |
|  | Fairly high or high level of support needed | 365 | 52.8% | 132 | 34.5% | 76 | 26.0% | 33 | 17.9% |
|  | *Missing* | 20 |  | 1 |  | 3 |  | 1 |  |
| Loneliness and social isolation | No or little support needed | 200 | 28.9% | 185 | 48.2% | 168 | 57.7% | 117 | 64.6% |
|  | Moderate level of support needed | 132 | 19.1% | 79 | 20.6% | 67 | 23.0% | 38 | 21.0% |
|  | Fairly high or high level of support needed | 359 | 52.0% | 120 | 31.3% | 56 | 19.2% | 26 | 14.4% |
|  | *Missing* | 20 |  | 0 |  | 4 |  | 4 |  |
| Regaining sense of purpose and meaning in life | No or little support needed | 219 | 31.7% | 177 | 46.2% | 176 | 60.5% | 124 | 67.4% |
|  | Moderate level of support needed | 149 | 21.6% | 94 | 24.5% | 56 | 19.2% | 32 | 17.4% |
|  | Fairly high or high level of support needed | 323 | 46.7% | 112 | 29.2% | 59 | 20.3% | 28 | 15.2% |
|  | *Missing* | 20 |  | 1 |  | 4 |  | 1 |  |
| Finding balance between grieving and other areas of life | No or little support needed | 187 | 27.1% | 172 | 44.9% | 186 | 63.7% | 128 | 69.6% |
|  | Moderate level of support needed | 193 | 27.9% | 127 | 33.2% | 57 | 19.5% | 35 | 19.0% |
|  | Fairly high or high level of support needed | 311 | 45.0% | 84 | 21.9% | 49 | 16.8% | 21 | 11.4% |
|  | *Missing* | 20 |  | 1 |  | 3 |  | 1 |  |
| Managing and maintaining my relationships with friends and family | No or little support needed | 259 | 37.4% | 231 | 60.8% | 209 | 71.8% | 120 | 65.2% |
|  | Moderate level of support needed | 183 | 26.4% | 86 | 22.6% | 49 | 16.8% | 40 | 21.7% |
|  | Fairly high or high level of support needed | 251 | 36.2% | 63 | 16.6% | 33 | 11.3% | 24 | 13.0% |
|  | *Missing* | 18 |  | 4 |  | 4 |  | 1 |  |
| Participating in work, leisure or other regular activities | No or little support needed | 289 | 42.2% | 229 | 59.9% | 222 | 76.0% | 139 | 76.0% |
|  | Moderate level of support needed | 164 | 23.9% | 86 | 22.5% | 37 | 12.7% | 27 | 14.8% |
|  | Fairly high or high level of support needed | 232 | 33.9% | 67 | 17.5% | 33 | 11.3% | 17 | 9.3% |
|  | *Missing* | 26 |  | 2 |  | 3 |  | 2 |  |
| **Practical support** |  |  |  |  |  |  |  |  |  |
| Getting relevant information and advice | No or little support needed | 365 | 53.3% | 302 | 78.6% | 256 | 87.1% | 161 | 87.0% |
|  | Moderate level of support needed | 153 | 22.3% | 46 | 12.0% | 24 | 8.2% | 14 | 7.6% |
|  | Fairly high or high level of support needed | 167 | 24.4% | 36 | 9.4% | 14 | 4.8% | 10 | 5.4% |
|  | *Missing* | 26 |  | 0 |  | 1 |  | 0 |  |
| Practical tasks | No or little support needed | 378 | 54.8% | 303 | 78.9% | 261 | 88.8% | 158 | 85.4% |
|  | Moderate level of support needed | 150 | 21.7% | 48 | 12.5% | 17 | 5.8% | 19 | 10.3% |
|  | Fairly high or high level of support needed | 162 | 23.5% | 33 | 8.6% | 16 | 5.4% | 8 | 4.3% |
|  | *Missing* | 21 |  | 0 |  | 1 |  | 0 |  |
| Looking after myself/family | No or little support needed | 433 | 62.0% | 307 | 79.9% | 245 | 83.3% | 159 | 85.9% |
|  | Moderate level of support needed | 159 | 22.8% | 45 | 11.7% | 28 | 9.5% | 16 | 8.6% |
|  | Fairly high or high level of support needed | 106 | 15.2% | 32 | 8.3% | 21 | 7.1% | 10 | 5.4% |
|  | *Missing* | 13 |  | 0 |  | 1 |  | 0 |  |

However, for participants meeting the threshold for indicated PGD, support needs remained high across all survey time points (see Fig. 1 and Supplementary File 5). For example, at the final survey time point (25-months post-bereavement, T4), 44.2% of respondents with indicated PGD continued to express high needs for support with coming to terms with how their loved one died (compared to 8.3% among those without indicated PGD), expressing their feelings and feeling understood (44.2% vs. 4.6%), loneliness and isolation (41.2% vs. 3.8%) and regaining a sense of purpose and meaning (40.4% vs. 5.3%).

**Fig 1:**
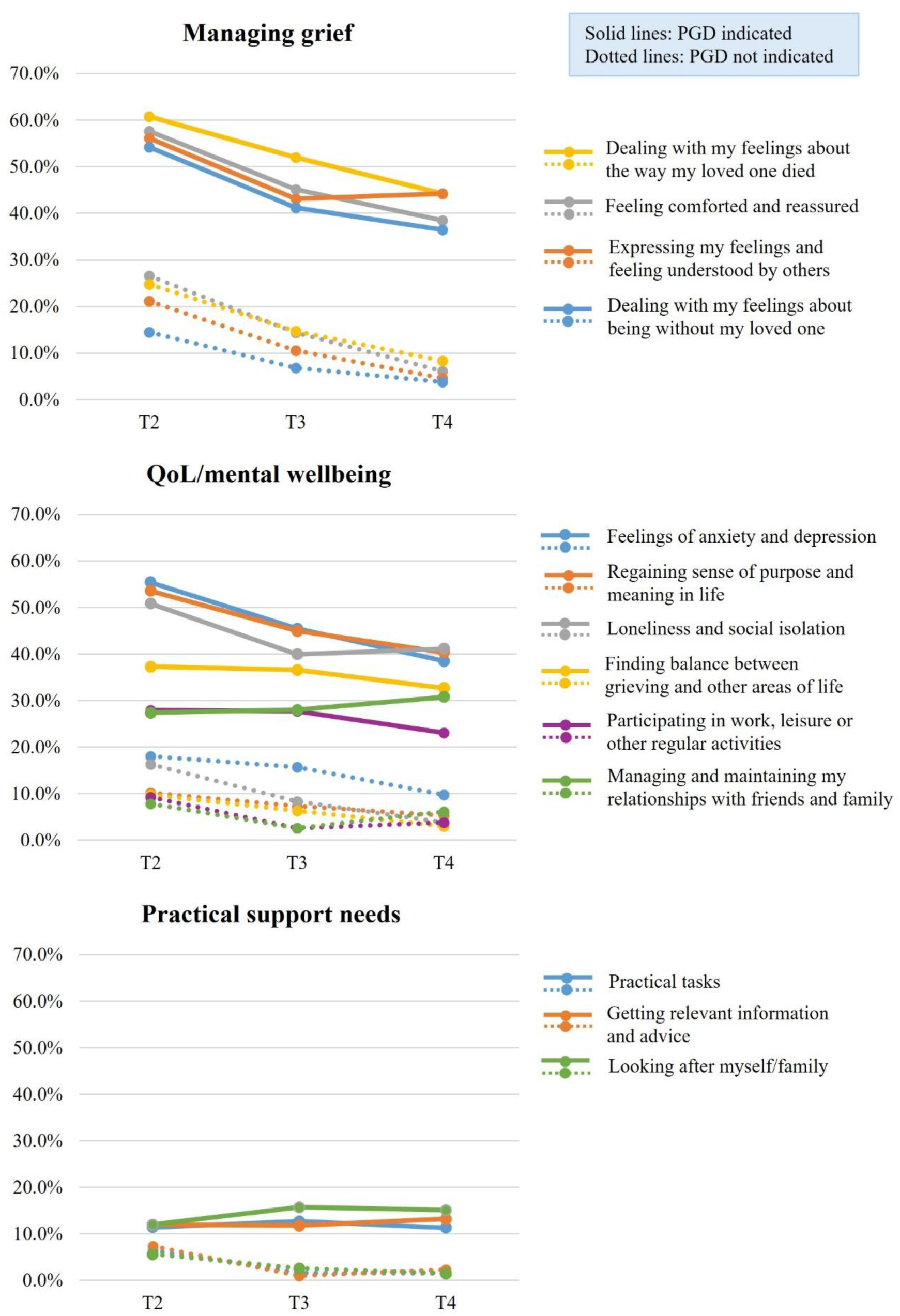
Levels of emotional and practical support needs at T2, T3 and T4 for participants with and without indicated PGD (T2-T4 = 8-, 13- and 25-months post-bereavement, respectively). TGI-SR measure was not used at baseline as PGD cannot be assessed before 6 months after death (39,40).

High-level practical support needs were less frequently reported, with about one in four respondents at T1 indicating that they needed fairly high or high levels of support with obtaining relevant information and advice (24.4%) and practical/administrative tasks (23.5%) (see Table 3). Though reports of practical support needs at the later time points were far less common in the overall sample (T3: 4.8%-7.1%; T4: 4.3%-5.4%), a minority of those with indicated PGD experienced high-level needs for help with practical matters (T3: 11.8%-15.7%; T4: 11.3%-15.1%) at these intervals (see Fig. 1 and Supplementary File 5).

Free text responses broadly aligned with the quantitatively assessed support needs, providing more contextualised insights into how these manifested in people’s individual bereavement experiences. Participants described practical challenges with the unfamiliar administrative tasks following their bereavement and expressed needs relating to managing their grief, in particular understanding and processing complex feelings relating to the circumstances of the death (including needing ‘answers’), talking about their experiences, coming to terms with their loss and resuming life without the person who died. They also described broader ‘wellbeing’ needs relating to perceived loss of purpose, dealing with loneliness and isolation, pandemic-related anxiety, and distress at being unable to support other family members due to social-contact restrictions, as well as anger and upset caused by dismissive public and political attitudes and behaviours towards the pandemic and pandemic-bereavement.

> *“I need support with understanding my grief reaction. Which doesn’t feel normal or healthy. Also the support needs to be trauma specialised due to the circumstances of my mother’s death from COVID and the general impact of the pandemic on every aspect of her death and circumstances after.”* (PID 020, bereaved daughter at T2)

Participants emphasised their need to connect with others with loss experiences similar to theirs (e.g. deaths due to Covid or in pandemic circumstances), highlighting the need for shared understanding and reassurance that they were not alone in their experiences. Many also expressed the need for professional bereavement or mental health support to deal with grief, depression, anxiety, trauma, PTSD symptoms and suicidal thoughts, while some also noted the need for support for and/or with other grieving family members, including children.

> *“I do feel that my son (who has always been anxious and sensitive), would probably benefit from more help which I don’t feel able to give.”* (PID 457, bereaved wife at T3)

### 3.3. Support use

People engaged with a range of support sources to help them cope with their bereavement and grief (see Table 4). Family and friends were by far the most used source of support, as captured both in the initial T1 survey (87.3%; n=621/711) and at T4 (90.0%; n=153/170). Among the six ‘non-family’ bereavement-specific support sources, the most commonly used types of support at all four time points were online groups/forums and one-to-one support, usually in the form of individual counselling. At baseline, about one in four respondents (28.6%; n=203/711) accessed online communities for support to help them cope with their bereavement. One in five (20.8%; n=148/711) accessed one-to-one support. Overall, the use of support tended to slowly decrease over the first year after the death (see Table 4). However, at T4, use of some of the informal or community-based support sources slightly increased again, as compared to T3, most notably the use of online community and telephone helpline/instant webchat support. More than a third of participants at T4 (33.0%; n=61/185) also reported that they had used written or audio-based resources (e.g. books, websites or podcasts on bereavement) in recent months.

Across all time points, participants with indicated PGD appeared to use all formal and informal support resources outside of family and friends to a greater extent than those without indicated PGD (e.g. at T3 29.4% vs. 10.4% % for online community, 27.5% vs. 16.6% for one-to-one support; see Fig. 2).

**Table 4:**
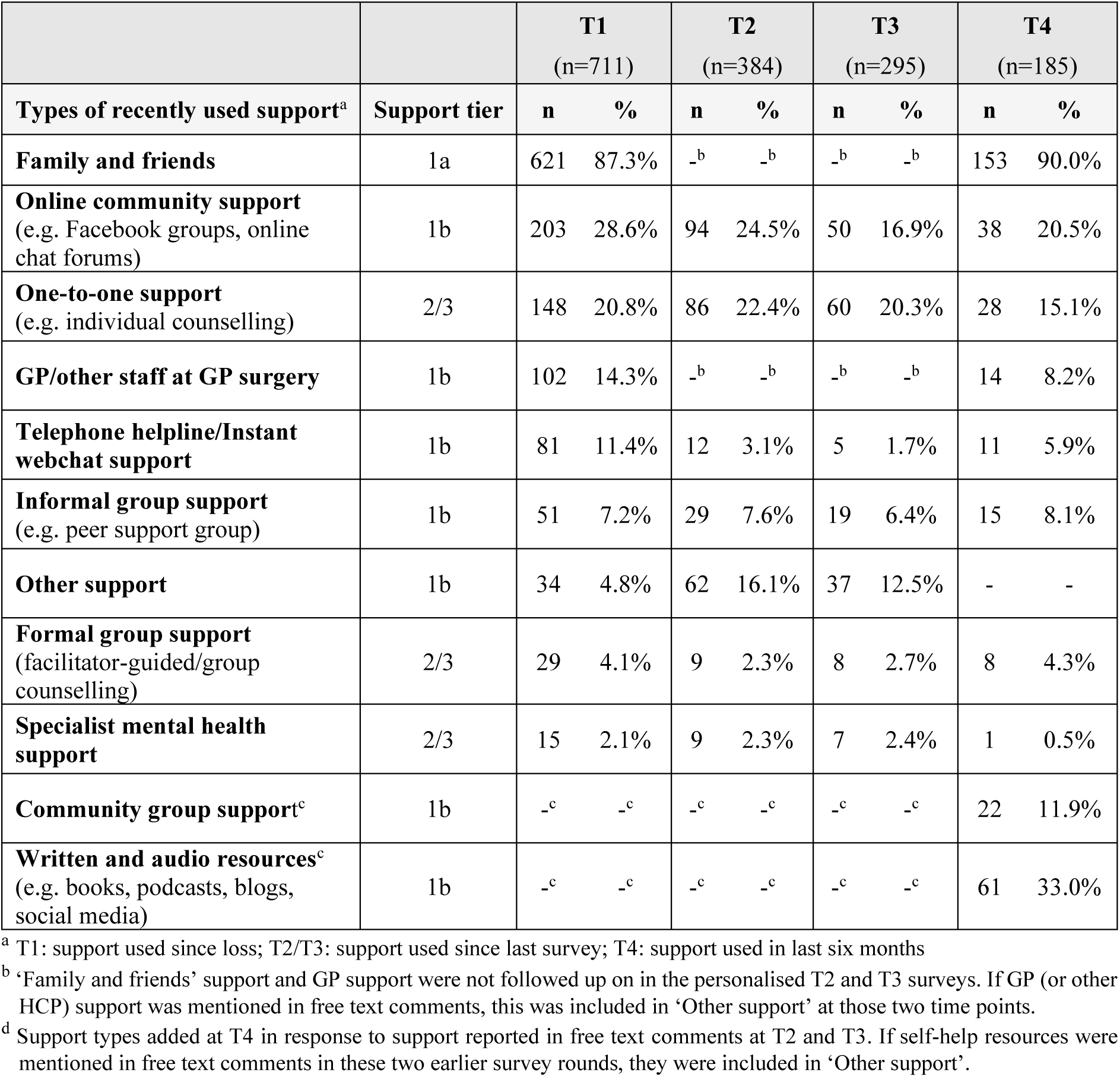
Informal and formal sources of bereavement-specific support used at the four time points (according to the three tiers of the Public Health Model. (**7**) **and ranked by % used at T1).**

|  |  | **T1**<br>(n=711) |  | **T2**<br>(n=384) |  | **T3**<br>(n=295) |  | **T4**<br>(n=185) |  |
| --- | --- | --- | --- | --- | --- | --- | --- | --- | --- |
| **Types of recently used support<sup>a</sup>** | **Support tier** | **n** | **%** | **n** | **%** | **n** | **%** | **n** | **%** |
| **Family and friends** | 1a | 621 | 87.3% | -<sup>b</sup> | -<sup>b</sup> | -<sup>b</sup> | -<sup>b</sup> | 153 | 90.0% |
| **Online community support**<br>(e.g. Facebook groups, online chat forums) | 1b | 203 | 28.6% | 94 | 24.5% | 50 | 16.9% | 38 | 20.5% |
| **One-to-one support**<br>(e.g. individual counselling) | 2/3 | 148 | 20.8% | 86 | 22.4% | 60 | 20.3% | 28 | 15.1% |
| **GP/other staff at GP surgery** | 1b | 102 | 14.3% | -<sup>b</sup> | -<sup>b</sup> | -<sup>b</sup> | -<sup>b</sup> | 14 | 8.2% |
| **Telephone helpline/Instant webchat support** | 1b | 81 | 11.4% | 12 | 3.1% | 5 | 1.7% | 11 | 5.9% |
| **Informal group support**<br>(e.g. peer support group) | 1b | 51 | 7.2% | 29 | 7.6% | 19 | 6.4% | 15 | 8.1% |
| **Other support** | 1b | 34 | 4.8% | 62 | 16.1% | 37 | 12.5% | - | - |
| **Formal group support**<br>(facilitator-guided/group counselling) | 2/3 | 29 | 4.1% | 9 | 2.3% | 8 | 2.7% | 8 | 4.3% |
| **Specialist mental health support** | 2/3 | 15 | 2.1% | 9 | 2.3% | 7 | 2.4% | 1 | 0.5% |
| **Community group support<sup>c</sup>** | 1b | -<sup>c</sup> | -<sup>c</sup> | -<sup>c</sup> | -<sup>c</sup> | -<sup>c</sup> | -<sup>c</sup> | 22 | 11.9% |
| **Written and audio resources<sup>c</sup>**<br>(e.g. books, podcasts, blogs, social media) | 1b | -<sup>c</sup> | -<sup>c</sup> | -<sup>c</sup> | -<sup>c</sup> | -<sup>c</sup> | -<sup>c</sup> | 61 | 33.0% |

<sup>a</sup> T1: support used since loss; T2/T3: support used since last survey; T4: support used in last six months
<sup>b</sup> 'Family and friends' support and GP support were not followed up on in the personalised T2 and T3 surveys. If GP (or other HCP) support was mentioned in free text comments, this was included in 'Other support' at those two time points.
<sup>d</sup> Support types added at T4 in response to support reported in free text comments at T2 and T3. If self-help resources were mentioned in free text comments in these two earlier survey rounds, they were included in 'Other support'.

**Fig. 2:**
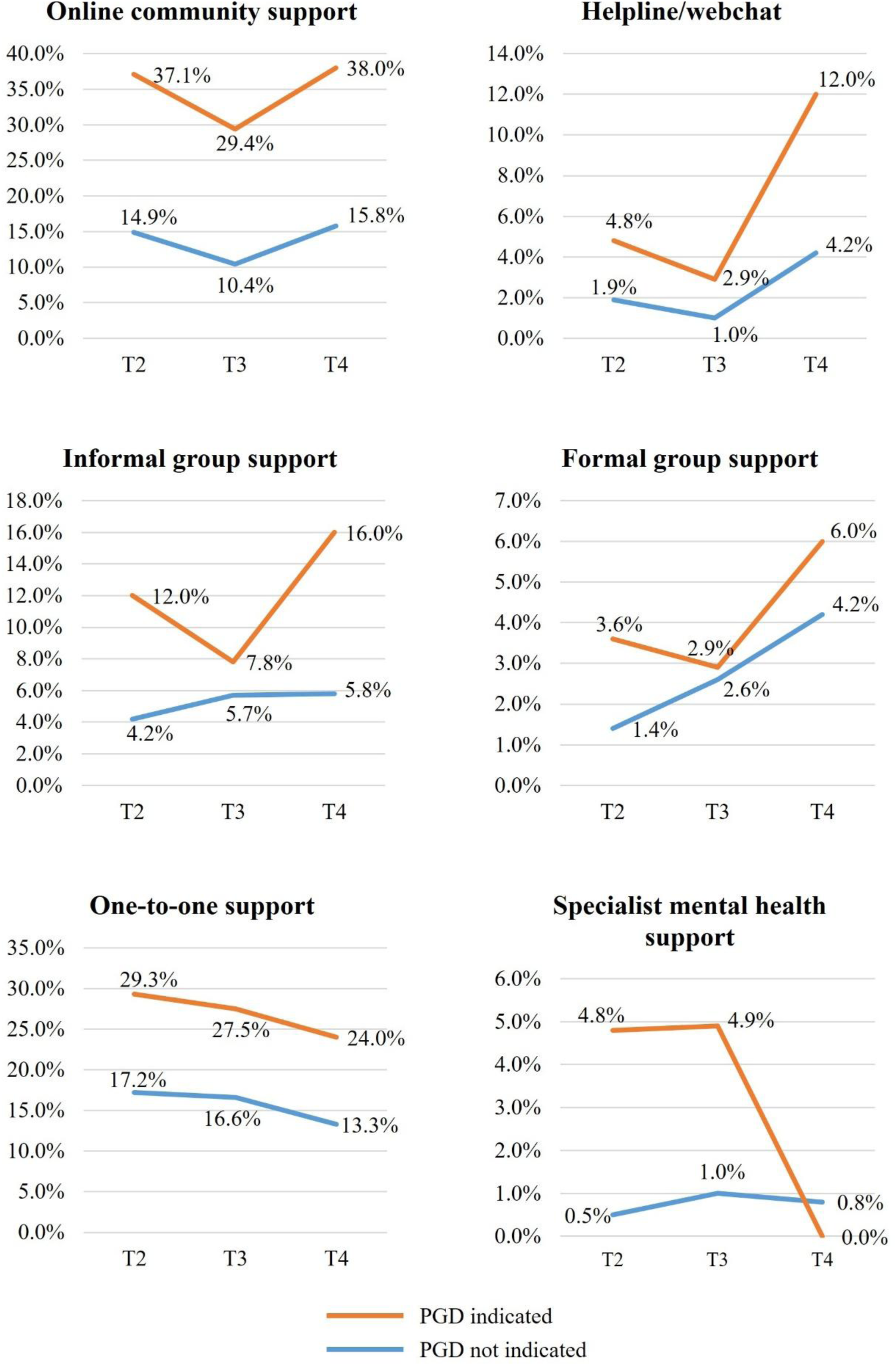
Informal and formal sources of support (outside of family and friends) used by participants with and without indicated PGD at T2, T3 and T4 (% used).

When looked at cumulatively and by highest accessed tiers of support at T2 and T3 (see Fig. 3), results show that the proportions of people who accessed top tier formal support (i.e. Tier 2/3: counselling or mental health support) increased within the first year. However, there was a substantial proportion of people (35.3%) with indicated PGD at T3 who had not been accessing this type of support during this time. At T4, 73.6% of this group had also not accessed formal support within the last 6 months, suggesting significant unmet need.

**Fig. 3:**
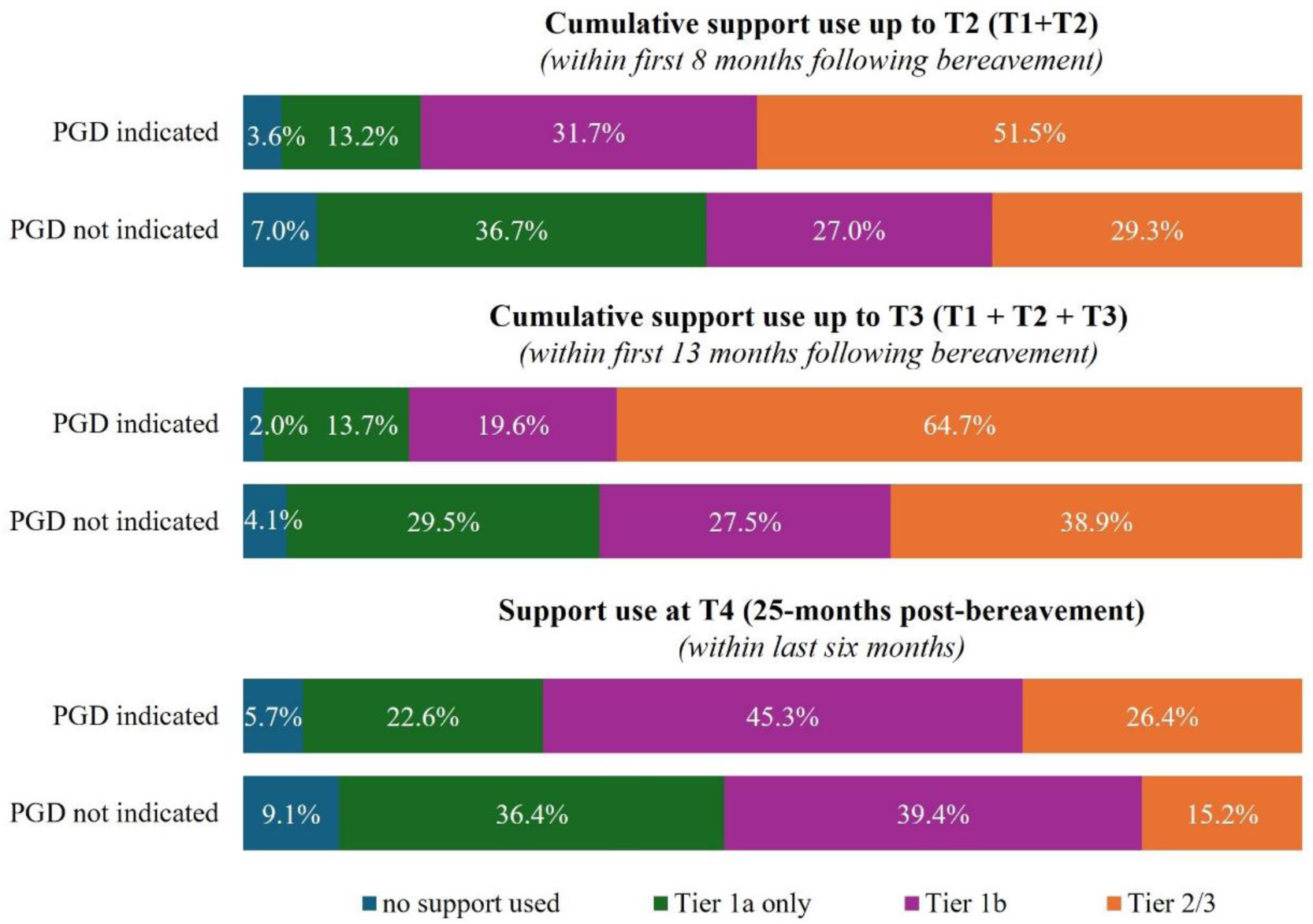
Highest level of support accessed by PGD status within the first and second year following the bereavement, using the three tiers of the Public Health Model (7).

### 3.4. Perceived impact of support

In the captured free text data, we identified eight major themes related to how people felt helped by the different types of support they had used following their bereavement, mapped onto the support domains of ‘Practical support’, ‘Managing grief’ and ‘Quality of life/mental wellbeing’ (see Table 5 and Supplementary File 6, with illustrative quotes).

**Table 5:**
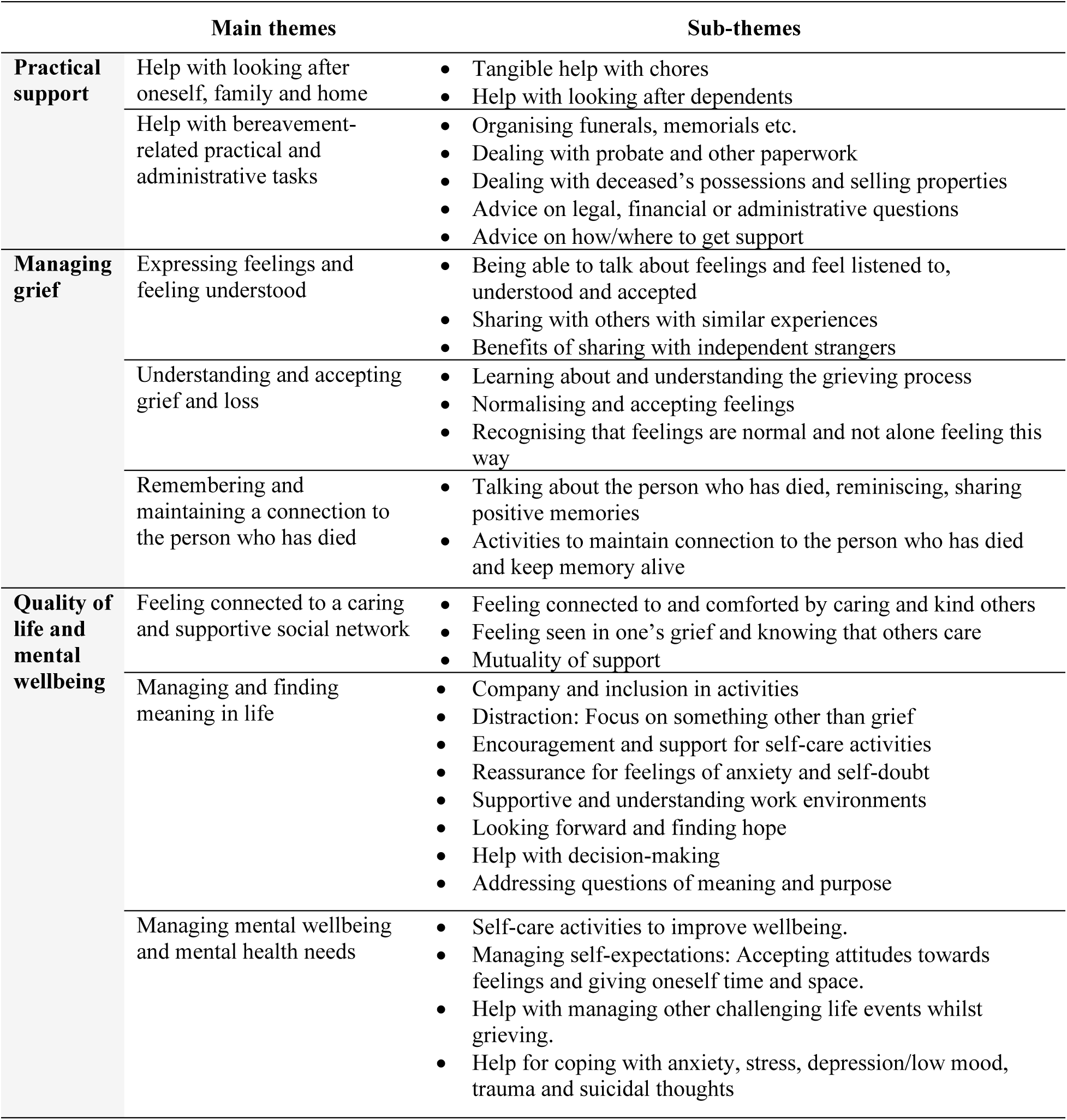
Overview of qualitative themes (mapped onto the three support domains ‘Practical support’, ‘Managing grief’ and ‘Quality of life/mental wellbeing’)

The narrative below captures how the impact themes were reflected in the different types of support people accessed, as well as activities and attitudes that people found helpful but were not explicitly linked to a specific external source of support. The majority of the free text responses analysed referred to support from family and friends, online community support and one-to-one support, which were the most frequently used sources of support. Almost all themes were reflected across the different time points in relation to specific support types; where differences occurred these are noted in the narrative.

#### 3.4.1 Practical support

Family, friends and sometimes neighbours were the primary source of practical help mentioned at the three time points within the first year following the bereavement (T1, T2 and T3). Participants appreciated being helped with household chores, meals and shopping, looking after children and pets, transport and financial issues, sometimes mentioning how a lack of focus, motivation or energy made such tasks difficult for them. In addition, people described being helped with bereavement-related administrative tasks relating to organising funerals/memorials, settling the deceased’s affairs (e.g. probate, paperwork) and help with/advice on handling legal and financial matters. Some reported obtaining professional help, e.g. from solicitors to tackle probate and property sales. Online groups or forums were occasionally mentioned as sources of informational support in the form of advice on where/how to get support (e.g. counselling, financial aid).

> *“My older sister has done the vast majority of clearing Mum’s house and sorting paperwork because I have become quickly overwhelmed by it.”* (PID 523, bereaved daughter at T2)

#### 3.4.2 Managing and coping with grief

How people felt helped with processing their loss and managing their feelings centred around the themes of ‘Expressing feelings and feeling understood’, ‘Understanding and accepting grief’ and ‘Remembering and maintaining a connection to the person who had died’.

##### Expressing feelings and feeling understood

The benefits of talking about grief and feelings without fear of judgment and feeling listened to and understood were experienced in relation to all support types. People described how helpful they found being able to talk openly about their experience with trusted family members and friends, especially due to the perceived sense of mutual support in their shared grief. Some experienced particular benefit from talking to close others who had previous bereavement experiences similar to theirs (e.g. traumatic or sudden death; parental loss).

> *“Friends who have lost someone. They understand where I am 2 years later.”* (PID 106, bereaved daughter at T4)

Online forums and groups provided an easily accessible platform/opportunity for many to share their experiences in the form of written comments with a wider group of bereaved others not directly connected to them. Feeling understood, validated and reassured by others with similar experiences brought great comfort. Likewise, people derived very similar benefits from talking, sharing and empathising with fellow bereaved persons in informal and formal bereavement support groups, which due to pandemic restrictions were usually conducted via group video calls.

> *“The [online] forum has been a good outlet. I’m not good at talking about my feelings or thoughts, but putting it on the forum and receiving responses from people who know exactly how I feel is comforting. Even just reading what others are going through helps to know I’m not alone.”* (PID 629, bereaved daughter at T1)

Individual counselling, usually via phone or video call, provided a dedicated space and time to reflect on the experience of loss in a non-judgemental environment and to safely connect with, explore, unpick and understand complex feelings (e.g. guilt, regret and anger; relationship to deceased person; changing family dynamics). People highlighted how talking to an independent third person enabled them to ‘take their mask off’ (PID 739 at T2) and openly share thoughts they worried may otherwise upset friends and family. At the same time, some felt that learning to talk about their grief in this context also helped them initiate conversations with their family and friends.

> *“Weekly counselling gives me an anchor to my week. A safe space to connect with challenging feelings, say things some people might find hard to listen to. She wasn’t emotionally connected to my partner so I don’t have to take care of her grief. That is greatly valued.”* (PID 484, bereaved wife at T2)

Some also appreciated that counselling provided them with a space to keep talking about their grief without the perceived social expectations and pressures of ‘needing to have moved on’, while also feeling encouraged to explore different perspectives.

> *“Counselling - she is a very skilled counsellor and I feel safe with her. She understands the grief is a much longer process than most other people. It means I don’t feel embarrassed explaining how lonely and desperate I still feel. Other people would not expect me to say those things as it is over two years.”* (PID 052, bereaved wife at T4)

##### Understanding and accepting grief and loss

Counselling in particular helped people understand the grieving process and recognise that their thoughts, feelings and ways of coping were normal. This provided reassurance that they were coping and helped them to become more accepting towards their grief journey/experience. Helplines and instant webchat services were used less frequently in this cohort but also enabled people to disclose their grief to a non-judgemental, independent person and to be reassured that what they were experiencing was normal. This support was often ‘one-off’ and was sought at times when people needed a listening ear but didn’t know where else to turn/felt they needed to reach out somewhere.

> *“bereavement line that is open 24/7 - just able to listen when you need it*.” (PID 513, bereaved wife at T2)

Connecting with others with shared experiences in online communities and bereavement support groups also helped people to understand more about their own feelings and experiences, enabling them to realise that they are not alone in feeling the way that they do and to recognise these as ‘normal’ reactions to their bereavement, and specifically pandemic bereavement. People found it reassuring to learn how others were feeling and coping and realise that their own emotions were very similar (‘I’m not the only one.’).

> *“It has been invaluable being put in contact with other people who have lost their parents to Covid. Covid grief feels so different and I feel many counsellors don’t know how to deal with it. I have a zoom call once a week with a group of ladies who have lost their partners or dads to Covid. It helps so much. It makes me feel not alone. It makes me feel like I can deal with another week and that I’m not crazy.”* (PID 710, bereaved daughter at T1)
>
> *“Just talking to people who understand and listening, and realising these feelings are normal.”* (PID 709, bereaved wife at T3)

In addition, people took to self-help resources such as grief books, bereavement-related websites and social media channels, podcasts, webinars and online grief events to understand more about grief and reflect on their own experiences. Such resources again helped people to feel reassured by learning about and relating to others’ similar experiences.

> *“It [podcast] has been something I can listen to while out walking and hear other people’s experiences. Helps me to process my own feelings and recognise that what I’m feeling is normal.”* (PID 070, bereaved daughter at T1)

People also described how keeping their focus on positive aspects of their bereavement – such as death having brought an end to suffering or ‘good death’ experiences – helped them to accept the death and their loss.

##### Remembering and maintaining a connection to the person who has died

Being able to talk about the person who had died and share positive memories was another important aspect of support that helped people to manage their grief. It was felt to be a crucial part of conversations with those who had also been close to the deceased person, alongside other activities, undertaken either with others or by themselves, to maintain a connection to the deceased person such as visiting their grave, maintaining shared activities or interests or writing to/about the deceased person. Some people also valued being able to share their memories in online communities, bereavement support groups and with counsellors.

> *“My children have allowed me to talk about my Mum (their Nana) and we are now able to start remembering the happy times and not just the trauma of her passing.”* (PID 178, bereaved daughter at T3)

#### 3.4.3 Quality of life and mental wellbeing

The majority of comments in this domain related to the importance of social support for comfort and wellbeing and people’s ability to resume activities, cope with everyday life and move forward after bereavement.

##### Feeling connected to a caring and supportive social network

The importance of continuous emotional support and having a sense of connectedness to a caring and supportive network dominated people’s accounts of how they felt helped at an emotional and social level and re-adapted to everyday life after their bereavement. Knowing that others care was key: people felt greatly comforted by family and friends acknowledging their loss, showing them kindness and empathy and communicating that they, regardless of long how ago the bereavement occurred, continued to be available for support, e.g. by continuing to check in, inviting conversations and being prepared to listen while also accepting people’s need for time and space to grieve.

> *“Friends coming to me of their own accord and making sure I know they care has helped too.”* (PID 057, bereaved daughter at T2)

Online groups/forums as well as peer support groups enabled people to feel particularly connected to a kind and caring community of others ‘who get it’ due to their shared experience of loss. They found meaning in supporting each other and, most importantly, felt less alone and isolated. Some forged new friendships online and group members often remained in contact after/outside of organised meetings.

> *“I was able to express my emotions and feelings at that time with people who had gone through the bereavement process themselves. […] It also helped that I could offer some support however small to other people going through what I was going through.”* (PID 584, bereaved son at T1)

##### Managing and finding meaning in life

Support from others was also important with regards to resuming activities and responsibilities, helping participants to find balance between grieving and managing other aspects of life. People greatly appreciated the company of family and friends and being encouraged to and included in activities that involved getting out of the house and being with others (e.g. socially distanced walks). This provided distraction, an opportunity to focus on something other than their grief and generally helped them to feel better and less lonely. Some also valued the reassurance and motivation/encouragement given by family and friends to help them tackle tasks and take care of themselves. Although many people experienced difficulties at work, supportive work environments were greatly appreciated and helped people return to and cope at work. Positive experiences included considerate line managers and understanding colleagues looking out for the bereaved person, flexibility around adjustments to work responsibilities and hours (e.g. phased returns to work), and opportunities to talk and share. Work was sometimes highlighted as providing structure, focus and a sense of normality.

> *“I am going for walks and coffee (socially distance of course!) with friends too, has been good just to talk about stuff, not just about my husband and the loss, but holidays and hope too. I have found friends and family just being there, and knowing there are people I could call at any time of day or night if I needed to has been probably the best help.”* (PID 284, bereaved wife at T1)

Talking about the future and making plans with friends and family helped people to look ahead. Online communities and support groups also provided reassurance and hope for the future amidst grief, with people taking comfort from observing the coping and progress made by others.

> *“The [bereavement support charity for bereaved partners] site has helped me to see that hopefully I will come out the other end in one piece!”* (PID 380, bereaved wife at T2)

Counselling provided support with dealing with challenges in other areas of their life, such as the ongoing pandemic, work stress and caring responsibilities, managing relationships with friends and family, as well as the wider implications of their bereavement. It also helped with making decisions and provided a space to address wider questions around meaning and purpose brought on by the bereavement. Some highlighted how they felt encouraged by the perceived progress they made in their counselling and grief journeys.

> *“The bereavement counselling has helped me to talk to someone not emotionally involved in my dad’s death and talk through some of the anger and frustration and sadness. Also how that has impacted my personal relationships, mental health and work. Having the sessions helped me decide to contact my GP and take a break from work.”* (PID 199, bereaved daughter at T1)

##### Managing mental wellbeing and mental health needs

Participants highlighted a range of different ways in which they looked after their emotional and mental wellbeing during this difficult time, involving both self-initiated self-care activities and receiving support from others. Journaling, being in nature, mindfulness apps/courses, exercising, eating well and getting enough sleep, time alone, and keeping busy were some of the most frequently mentioned self-care strategies perceived as helpful.

Some expressed how approaching all and any feelings with an accepting attitude and giving oneself time and space to feel, learn about and express feelings helped them be kinder to themselves and maintain a more positive outlook. Appreciating life more after experiencing a death brought comfort and helped them come to terms with their loss, while others found strength in their faith or spirituality.

> *“Allowing myself time to grieve-it comes in waves. Sometimes I just sit on the floor and cry and cry, and I’ve learnt to let myself do that.”* (PID 105, bereaved granddaughter at T2)

Counselling helped some with tackling negative thoughts and managing self-expectations and offered suggestions for coping strategies, including expressive and self-care activities (e.g. art, journaling, exercise). Friends and family also provided reassurance to mitigate anxiety, low self-confidence and doubts about whether they were coping.

> *“I have needed reassurance from people who know me and my counsellor that I am OK and coping. It has felt hard to assess this for myself at times. Grief can cause a lot of self doubt and lockdown has not helped as isolation means we receive less ‘evidence’ that we can do things.”* (PID 535, bereaved friend at T2)

For some, counselling and specialist mental health support was also vital for coping with more severe mental health problems such as anxiety, depression/low mood, trauma/post-traumatic stress and suicidal thoughts. GPs were sometimes approached about medication to help with depression, anxiety and/or sleep, counselling referrals and sick notes.

#### 3.4.4 Unhelpful features of support

When reflecting on the support they had used/received, participants sometimes also described aspects that made support unhelpful or challenging, with comments mostly relating to family and friends and counselling.

Across all time points, many felt that their family and friends did not understand their bereavement and its (long-term) impact. Despite participants’ intense need to talk about their experiences and feelings, family and friends often avoided conversations for fear of not knowing what to say, used unhelpful platitudes in an attempt to comfort and reassure, provided unsolicited advice or tried to distract instead of just listening. Participants also described how support often quickly waned despite their ongoing need for support and that the perceived expectation to have gotten ‘over it’ soon left them unable to bring up their bereavement again.

> *“Friends and family seemed to have ‘moved on’ and there is little space to talk about my loss with them now.”* (PID 027, bereaved daughter at T4)

Negative experiences with counselling support were much less frequently described, but included poor rapport with the practitioner, a perceived lack of skill, knowledge or professionalism, feeling that needs or expectations were not being met, and experiencing the counselling process as too upsetting or emotionally draining. Some expressed that support providers lacked sufficient understanding of the unique challenges associated with bereavement during a pandemic while a small number also described a perceived lack of sensitivity for religious or cultural needs. Especially at T1 and T2, some commented that they would have preferred counselling support in-person rather than remotely via telephone or video call due to the pandemic restrictions.

> *“I lost faith in bereavement services after I had a few sessions with [bereavement charity] and the counsellor was eating on the phone, it made me feel unimportant and he also kept talking over me and guessing what I was going to say instead of letting me talk, so that has massively put me off approaching those services.”* (PID 503, bereaved friend at T4)

Fewer comments related to unhelpful support experiences in online community groups or forums. Those that were expressed included difficulties relating to other people’s (different) grief experiences, feeling discouraged or upset by others’ grief or their negative outlook and unkind comments. Group-based support was also experienced as less helpful if the group members were too different in their experiences (e.g. regarding the type of their bereavement or where they are at in their bereavement journey), with some also commenting on the IT challenges related to group video calls.

### 3.5. Future support preferences

The three most preferred support sources if participants were to be bereaved again in the future, assessed in the last survey round (T4), were family and friends (96%), in-person one-to-one support (75%) and self-help resources such as books and podcasts (63%), followed by support from GPs (61%), telephone/video-based one-to-one support (58%), telephone helplines (58%) and formal in-person bereavement support groups (57%) (see Fig. 4). Preferences were most mixed with regards to all forms of online group support (online meetings of informal peer-support groups, community groups or formal bereavement support groups) and telephone/video-call based specialist mental health support, with similar proportions of participants expressing that they would strongly/quite like or strongly/slightly dislike these kinds of support.

**Fig. 4:**
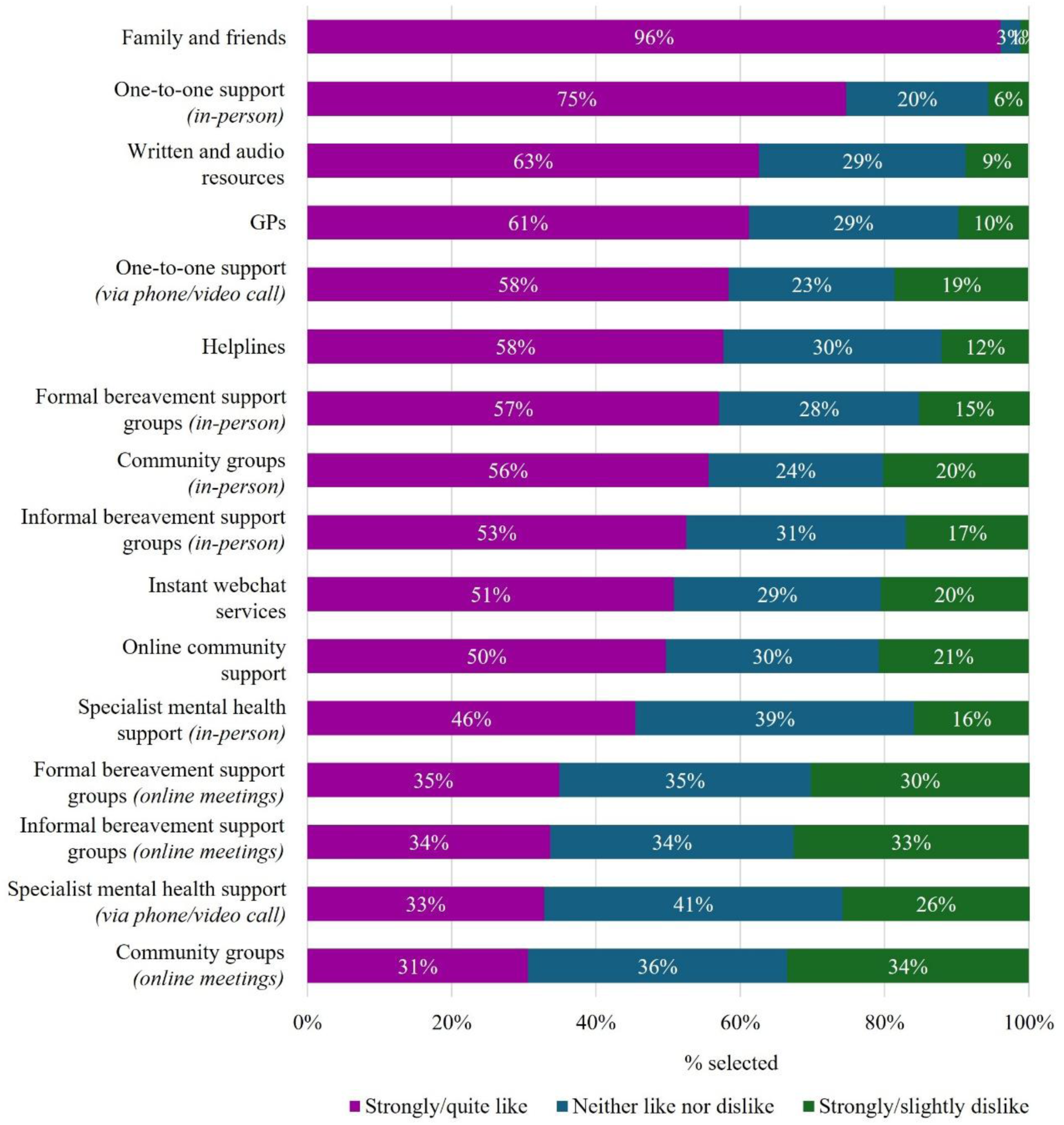
Support preferences for future bereavements under non-pandemic circumstances (captured at T4).

While the future support preferences of participants with indicated PGD at T4 were similar to those of participants without indicated PGD for many support options, there were also some marked differences (see Supplementary File 7). Those with indicated PGD more commonly preferred: online community support (70.0% compared to 41.5% without PGD), informal instant support sources like helplines (64.7% vs. 55.0%) and online webchat support (59.6% vs. 47.3%), and in-person formal and informal support groups (70% vs. 52% and 61.5% vs. 48.8%, respectively). Those with indicated PGD also more frequently expressed a preference for specialist mental health support, especially in-person (60.0% vs. 39.7%; remote support: 41.7% vs. 29.4%), in proportions notably higher than those who accessed such support in this cohort (< 5%; see Fig. 2).

## 4. Discussion

This research provides new evidence on changing patterns of bereavement support needs, support use and support experiences amongst a cohort of participants bereaved during the Covid-19 pandemic. Findings demonstrate high levels of support needs across multiple domains relating to managing grief and quality of life/wellbeing in the earlier months of bereavement, which for people with indicated PGD endured over time. Participant experiences of different types of informal and formal support demonstrate their many benefits and efficacy in meeting these needs. However, inadequacies in support were highlighted, particularly from friends and family, while a significant proportion of participants with indicated PGD missed out on any kind of formal bereavement or mental health support in the first year of their bereavement. These findings have important implications for policymakers, service providers and communities in terms of recognising and responding to the multiple, varied and sometimes enduring support needs of people bereaved in pandemic and non-pandemic times.

### 4.1. Support needs

In line with previous findings, we found that the highest support needs were emotional (rather than practical), and related to managing grief and feelings surrounding the loss, social isolation and connectedness (4,12,13) as well as feelings of anxiety and depression and regaining meaning and purpose. These findings provide empirical support for the relevance and comprehensiveness of the support need items included in our scale, and the two coping and wellbeing outcomes described in previous research (28,29), as well as related aspects of loss and restoration-orientated coping (30), including meaning-reconstruction (43). The severity and prevalence of these needs were very likely influenced by the unusual and distressing dying experiences, social isolation and lack of access to support caused by the pandemic (20,34–36,44–46), as also reflected in relatively high levels of indicated PGD in this cohort (33). As with indicated PGD levels (33), all support needs markedly decreased over the two-year study period, suggesting that most participants adjusted to their loss and with the easing of pandemic restrictions benefited from being able to reengage with everyday life activities and networks (44).

Among the substantial minority of participants who met the threshold for indicated PGD, however, these support needs remained high across survey-time points in the two-year follow up period, despite the return to more ‘normal times’-suggesting their continuing relevance within high-risk groups. While loss-oriented needs (30) relating to managing grief had slightly greater prevalence, more general wellbeing (and restoration-oriented) needs were also salient and keenly felt. Notably, ‘social’ needs relating to expressing feelings and feeling understood, loneliness and social isolation and managing relationships also increased slightly between one and two years post-bereavement in this group of participants. Loneliness (47) and lack of social support over time (33) have been associated with higher levels of prolonged grief symptoms, with PGD symptoms recently shown to predict social, emotional and general loneliness (48). Relatedly, severe grief reactions have been associated with more negative social reactions from others (49), with bereaved people with PGD shown to elicit a stronger desire for social distance than people with non-clinical grief reactions (50,51). It has been suggested that people with PGD may suppress their difficult grief-related emotions when with others and that the cognitive and emotional demands associated with this masking process motivate a preference for solitude and/or withdrawing from others (52). The strongly expressed ‘social’ needs of our PGD participants, alongside other restoration-oriented needs such as finding new meaning and purpose, arguably challenge assumptions that high-risk individuals favour solitude and withdrawal, or become fixated within a loss orientation at the expense of restoration orientations and ‘oscillation’ between the two (30). Findings therefore suggest important roles for friends, family and services in supporting both types of orientations in people struggling with prolonged, complex grief, including needs relating to social (as well as psychological) aspects of grief and coping, which are arguably under-recognised in the current diagnostic criteria for Prolonged Grief Disorder (e.g. (53,54).

### 4.2. Support experiences and preferences

*How* people felt helped by different types of support appeared similar to previous reports of helpful features of support (3,4,14,16,17,22,32), despite the shift to digital communication tools and remote forms of support provision during the pandemic. The different ways in which participants benefited from support closely reflected the described support needs for this cohort, and the two previously identified support outcomes and functions relating to coping and wellbeing (28,29), with variations seen across support types in terms of the more specific ways in which people benefited.

As in other studies (3,11–13), family and friends were the primary source of support, received by almost all participants across the different time points, and strongly favoured if bereaved again. Key benefits reflected practical support needs by means of tangible help with both everyday and bereavement-related practical and administrative tasks, especially in early bereavement (3,4,13–18). Emotional and social support that enabled expressions of grief, coping with feelings and maintaining social connection, purpose and distraction from grief was also valued. Supportive behaviours included the continued willingness to listen and talk about grief and the person who died, expressions of empathy, care and kindness, and efforts to offer company and inclusion in social activities (3,4,12–16). However, participants also commonly experienced issues and limitations with this support, including others lacking understanding of the impact of their bereavement, avoiding conversations or expecting them to ‘move on’, as also indicated in the high levels of unmet (and enduring) social support needs already described, and consistent with other research (3,4,12,13,16,17,19).

To some extent online community support (e.g. online forums/Facebook groups) and peer support groups appeared to compensate for these deficits, with around a quarter of participants using online communities, and much lower proportions using support groups - likely in part due to pandemic restrictions. Such support helped people to manage their grief by sharing with and feeling understood by others with similar experiences, normalising and making sense of their grief experiences and sometimes remembering those who had died, benefitting their wellbeing through feeling connected, less isolated and more positive and hopeful, in line with previous findings (32,55–57). Few negative experiences were reported, although some disliked or experienced difficulties with the online format of support groups. While the high levels of participation in online communities was likely due to the pandemic context and the limitations of other forms of support (20,25), its popularity as a future support source, alongside support groups, suggests the benefits of these forms of peer support going forwards for high and lower risk groups.

One-to-one support, such as counselling, was the third most commonly used form of support, and the second most popular form of future support, with the observed increases in pandemic-related complex grief (33) likely elevating needs for more formal support (25,58,59). Consistent with previous research, counselling helped to address many of the identified support needs, most commonly (and reflecting loss-orientation (30)) by providing a safe space to express and explore feelings (including those too difficult or upsetting to share with close others), learn about and understand the grieving process and recognise and accept their feelings and ways of coping as normal (22,32). Restoration-oriented benefits to wellbeing included support with managing relationships and other challenging areas of life, making decisions and finding meaning and purpose, as well as more general coping and self-care activities. For participants with more severe mental health needs, counselling and specialist mental health support were also vital to manage these. Although participant experiences were mostly positive, where difficulties existed these tended to be either due to lack of rapport/ competencies of individual counsellors (22), or unhappiness with virtual modes of delivery. Further, while participants with indicated PGD were overall more likely to engage with such support (8,60), worryingly many did not receive any formal counselling or mental health support, suggesting significant and enduring unmet needs. These findings are consistent with previous observations that bereaved individuals who are most in need of health care might not be receiving such help (61–63).

### 4.3. Implications for policy and practice

To a large extent, these findings provide support for public health models of bereavement support (7–10). At a universal level, they demonstrate the essential role of social networks, as well as the potential of self-help resources and self-directed approaches for managing grief, improving wellbeing, and assisting with practical tasks. However, the difficulties that people experienced with their networks also points to low levels of grief literacy (64), with people lacking the knowledge and skills needed to support others through grief. Public awareness and community initiatives are needed to help increase grief literacy (64) and understanding of the long-term and isolating impact of bereavement and the uniqueness and variability of grief (64) – including in its acute and prolonged forms – to foster more compassionate and supportive communities and reduce social isolation for bereaved people (64,65).

In line with public health models (7–10), findings also illustrate the benefits of group and one-to-one support for the smaller proportions of people who needed and accessed it, highlighting the specific benefits of counselling for addressing complex and difficult feelings, as well as the importance of specialist psychological support for those with mental health needs. However, the significant proportion of people with indicated PGD who did not appear to receive any formal support is of concern, highlighting the need for improved screening processes and effective needs assessment to identify bereaved individuals at risk and enable timely access to adequate specialist support (66). It also underscores the importance of normalising help-seeking in longer-term bereaved populations and those with persisting high-level needs, with proactive outreach and signposting to available support across community and healthcare settings.

Although in-person formats for group and one-to-one support were more popular, online formats were nonetheless acceptable for a considerable proportion of people, suggesting the value of services offering both types of support where possible, or if in-person provision is not viable (27). The varied support needs, and popularity and commonly described benefits of the many types of self-directed, group and one-to-one support across the study cohort similarly suggests the need for services to offer a range of support-options, including for higher risk groups who seem likely to also benefit from social and self-directed approaches, in addition to recommended psychological support. Making a broad range of informal and formal support types and delivery formats available is crucial for enabling bereavement support that is flexible and adaptable to individual needs and preferences. This includes ensuring long-term availability of support as needs can persist (or re-emerge) long after the initial bereavement period (29). Greater awareness of available (local) support options and their potential benefits is needed both among bereaved individuals and the professionals who support them, in particular GPs, whose popularity as a hypothetical source of bereavement support is also noteworthy, despite low levels of access (20,67,68).

### 4.4. Strengths, weaknesses and implications for future research

This longitudinal mixed-methods study offers nuanced insights into support needs following bereavement and patterns and perceptions of support use, including comparisons between participants with and without indicated PGD. Although the severity and prevalence of support needs and patterns of support use must be considered in the context of the challenging circumstances of the COVID-19 pandemic (20,34–36,45,46), the findings are nonetheless useful for informing improvements to bereavement support generally and specifically for those with more complex and persistent support needs.

The convenience sample of participants was self-selecting and potentially biased towards people engaged with online communities and support providers who helped promote the study. Men and people from ethnically diverse communities were under-represented, despite targeted recruitment efforts, and distribution of the survey primarily online may also have limited participation from more elderly individuals and others with restricted digital access. Conclusions about some support types (e.g. formal and informal support groups) must also be considered with caution due to the small numbers of participants accessing such supports, which further declined over time due to participant attrition.

More insights are needed into the socio-demographic factors that shape patterns of bereavement support use, support preferences, and perceived helpfulness of different support types, and ways in which support use impacts on bereavement outcomes. Future research should in particular focus on including groups that were underrepresented in the current study – such as men and bereaved individuals from ethnically diverse backgrounds – to build an evidence base for improvements to bereavement support that reflects diverse experiences and needs. Given the identified limitations in the support that people perceive from their networks, research exploring the experiences and challenges associated with supporting bereaved friends or family members is also recommended. This evidence will be important to incorporate when developing strategies to increase grief literacy (64) and help communities offer more effective and sustained support to bereaved individuals (64,65).

### 4.5. Conclusions

This study illustrates the multiple and varied emotional and social support needs of bereaved people, which for those with indicated PGD endured over time. It also demonstrates how different types of informal and formal support can help to address these needs, and in ways which broadly reflect previously described support outcomes and functions for bereavement services (28,29). However, expressed disappointment and dissatisfaction with regards to support from friends and family, and under-utilisation of formal bereavement or mental health support amongst high-risk groups also suggests significant unmet need and missed opportunities. It is important for policy makers and support providers to find ways of strengthening the support available to people within their networks and communities and enabling access to a wide variety of support, according to people’s needs and preferences.

## Supporting information

Supplementary File 1: Survey checklist

Supplementary File 2: Template baseline survey

Supplementary File 3: Template follow-up surveys at T2 and T3

Supplementary File 4: Template follow-up survey at T4

Supplementary File 5: Support needs

Supplementary File 6: Full themes table

Supplementary File 7: Future support preferences

## Data availability statement

The datasets presented in this study can be found in online repositories. The names of the repository/repositories and accession number(s) can be found at: UK Data Service via https://reshare.ukdataservice.ac.uk/855751.

## Ethics statement

This study received ethical approval from the Cardiff University School of Medicine Research Ethics Committee (SMREC 20/59) and was carried out in compliance with all relevant local legislation and institutional requirements. All participants provided written informed consent before taking part in this research.

## Author contributions

EH and LS designed the study, led the application for funding, and were co-principal investigators. SG, KB, ES, SS, ML and AP were members of the research team or the study advisory group and contributed to the design of the study and survey. SG was responsible for survey dissemination and data management and carried out the quantitative and qualitative data analysis for this paper, with support from EH, ES, KB and RO. SG and EH drafted the initial manuscript, with all authors contributing to its development and reading and approving the final version.

## Acknowledgements

We thank everyone who completed our survey(s) for sharing their experiences, as well as all the individuals and organizations who helped disseminate the initial baseline survey.

