## Supplementary File 1: Survey checklist for "Support needs, support use and perceived helpfulness of support in a cohort of people bereaved during the COVID-19 pandemic: Insights from a longitudinal survey"

**Supplementary File 1:** Checklist for Reporting Results of Internet E-Surveys (CHERRIES) (1)

| ***Item Category*** | ***Checklist Item*** | ***Explanation*** | ***Page Number*** |
|---|---|---|---|
| **Design** | Describe survey design | Describe target population, sample frame. Is the sample a convenience sample? (In "open" surveys this is most likely.) | See *Study procedure* and *Participants* on page 6. |
| **IRB (Institutional Review Board) approval and informed consent process** | IRB approval | Mention whether the study has been approved by an IRB. | See *Ethical approval* on page 7. |
| | Informed consent | Describe the informed consent process. Where were the participants told the length of time of the survey, which data were stored and where and for how long, who the investigator was, and the purpose of the study? | See study information in *Supplemental File 2* (baseline survey), pages 1-3: information on background and purpose of study; research team incl. contact details; length of survey; confidentiality; data storage, protection and usage; risks and benefits of participating; sign-posting information for support services should participants feel they need support during or after completing the survey. |
| | Data protection | If any personal information was collected or stored, describe what mechanisms were used to protect unauthorized access. | See *Supplemental File 2* (baseline survey), pages 2-3. Anonymised research data will be kept securely for 10 years after the study is completed In line with Cardiff University policies and General Data Protection Regulations (GDPR; EU 2016/679) and the Data Protection Act 2018 (DPA 2018). Contact details for follow-up surveys/study updates were securely destroyed after the last survey round (T4) and provision of a final study update to all participants who had requested updates. |

| | | | |
|---|---|---|---|
| **Development and pre-testing** | Development and testing | State how the survey was developed, including whether the usability and technical functionality of the electronic questionnaire had been tested before fielding the questionnaire. | See *Survey development and contents: Baseline survey and follow-up surveys* on page 5 which also provides a reference to the baseline paper with more detailed information on the survey development (Harrop et al. 2021). Before the baseline/follow-up surveys were disseminated, internal pilot testing was carried to ensure usability and technical functionality of the online questionnaires. |
| **Recruitment process and description of the sample having access to the questionnaire** | Open survey versus closed survey | An “open survey” is a survey open for each visitor of a site, while a closed survey is only open to a sample which the investigator knows (password-protected survey). | See pages 5 and 6. Baseline survey (T1) was an open survey disseminated to a convenience sample. For the follow-up surveys (T2, T3, T4), a personalised survey link with their unique study participant ID was emailed to those T1 respondents who had consented to being contacted with additional surveys. |
| | Contact mode | Indicate whether or not the initial contact with the potential participants was made on the Internet. (Investigators may also send out questionnaires by mail and allow for Web-based data entry.) | See page 6. Baseline survey was primarily disseminated online but hardcopy postal surveys were available on request for all survey rounds. |
| | Advertising the survey | How/where was the survey announced or advertised? Some examples are offline media (newspapers), or online (mailing lists – If yes, which ones?) or banner ads (Where were these banner ads posted and what did they look like?). It is important to know the wording of the announcement as it will heavily influence who chooses to participate. Ideally the survey announcement should be published as an appendix. | See page 6. Baseline survey was advertised via social and mainstream media, voluntary sector associations and bereavement support organisations (e.g. via social media, webpages, newsletters, online forums). The provided survey invitation email templates and survey flyers are available upon request. |
| **Survey administration** | Web/E-mail | State the type of e-survey (eg, one posted on a Web site, or one sent out through e- | See page 6. All surveys were administered via the JISC online survey platform (2). |

| | | | |
|---|---|---|---|
| | | mail). If it is an e-mail survey, were the responses entered manually into a database, or was there an automatic method for capturing responses? | |
| | Context | Describe the Web site (for mailing list/newsgroup) in which the survey was posted. What is the Web site about, who is visiting it, what are visitors normally looking for? Discuss to what degree the content of the Web site could pre-select the sample or influence the results. For example, a survey about vaccination on a anti-immunization Web site will have different results from a Web survey conducted on a government Web site | See survey advertising above. In addition, the baseline survey link was also posted onto a bespoke study-specific website with a memorable URL (covidbereavement.com). This information is not included in the manuscript as the study website is no longer active. |
| | Mandatory/voluntary | Was it a mandatory survey to be filled in by every visitor who wanted to enter the Web site, or was it a voluntary survey? | See page 6. Voluntary survey. |
| | Incentives | Were any incentives offered (eg, monetary, prizes, or non-monetary incentives such as an offer to provide the survey results)? | See page 6. Survey was non-incentivised. |
| | Time/Date | In what timeframe were the data collected? | See pages 6 and 7. |
| | Randomization of items or questionnaires | To prevent biases items can be randomized or alternated. | See page 5. Non-randomised open and closed questions. |
| | Adaptive questioning | Use adaptive questioning (certain items, or only conditionally displayed based on responses to other items) to reduce number and complexity of the questions. | There was only minimal adaptive questioning across all four survey rounds, mainly used to prompt specification for 'Other' responses or to skip irrelevant questions (e.g. child-related questions for participants without children). |
| | Number of Items | What was the number of questionnaire items per page? The number of items is an important factor for the completion rate. | Number of main questions (with additional follow-up questions where applicable):<br>Baseline survey T1: 37<br>Follow-up survey T2: 25 |

| | | | |
|---|---|---|---|
| | | | Follow-up survey T3: 24<br>Follow-up survey T4: 25 |
| | Number of screens (pages) | Over how many pages was the questionnaire distributed? The number of items is an important factor for the completion rate. | Information no longer available after a recent upgrade to the online survey platform JISC (2). |
| | Completeness check | It is technically possible to do consistency or completeness checks before the questionnaire is submitted. Was this done, and if "yes", how (usually JAVAScript)? An alternative is to check for completeness after the questionnaire has been submitted (and highlight mandatory items). If this has been done, it should be reported. All items should provide a non-response option such as "not applicable" or "rather not say", and selection of one response option should be enforced. | Except for the informed consent statement that participants had to agree to be able to proceed to the survey, all survey questions were voluntary. There were no automatic prompts or alerts to notify participants if they accidentally or intentionally skipped a question. |
| | Review step | State whether respondents were able to review and change their answers (eg, through a Back button or a Review step which displays a summary of the responses and asks the respondents if they are correct). | Participants were able to review and change their answers through a Back Button. |
| **Response rates** | Unique site visitor | If you provide view rates or participation rates, you need to define how you determined a unique visitor. There are different techniques available, based on IP addresses or cookies or both. | N/A |
| | View rate (Ratio of unique survey visitors/unique site visitors) | Requires counting unique visitors to the first page of the survey, divided by the number of unique site visitors (not page views!). It is not unusual to have view | N/A |

| | | | |
|---|---|---|---|
| | | rates of less than 0.1 % if the survey is voluntary. | |
| | Participation rate (Ratio of unique visitors who agreed to participate/unique first survey page visitors) | Count the unique number of people who filled in the first survey page (or agreed to participate, for example by checking a checkbox), divided by visitors who visit the first page of the survey (or the informed consents page, if present). This can also be called "recruitment" rate. | N/A |
| | Completion rate (Ratio of users who finished the survey/users who agreed to participate) | The number of people submitting the last questionnaire page, divided by the number of people who agreed to participate (or submitted the first survey page). This is only relevant if there is a separate "informed consent" page or if the survey goes over several pages. This is a measure for attrition. Note that "completion" can involve leaving questionnaire items blank. This is not a measure for how completely questionnaires were filled in. (If you need a measure for this, use the word "completeness rate".) | N/A |
| **Preventing multiple entries from the same individual** | Cookies used | Indicate whether cookies were used to assign a unique user identifier to each client computer. If so, mention the page on which the cookie was set and read, and how long the cookie was valid. Were duplicate entries avoided by preventing users access to the survey twice; or were duplicate database entries having the same user ID eliminated before analysis? In the latter case, which entries were kept for analysis (eg, the first entry or the most recent)? | See *Supplemental File 2* (baseline survey), page 3. No cookies were used. |

| | | | |
|---|---|---|---|
| | IP check | Indicate whether the IP address of the client computer was used to identify potential duplicate entries from the same user. If so, mention the period of time for which no two entries from the same IP address were allowed (eg, 24 hours). Were duplicate entries avoided by preventing users with the same IP address access to the survey twice; or were duplicate database entries having the same IP address within a given period of time eliminated before analysis? If the latter, which entries were kept for analysis (eg, the first entry or the most recent)? | To protect participant anonymity, no IP addresses were recorded. |
| | Log file analysis | Indicate whether other techniques to analyze the log file for identification of multiple entries were used. If so, please describe. | No other measures were taken to prevent multiple entries from the same individual. However, as described in more detail in Harrop et al. 2021 ((3); see reference in *Sample characteristics* on page 7), a small number of baseline surveys (n=12) were identified as duplicates based on similarities in the provided contact and demographic information. For these participants, the first completed survey was retained for analysis. |

| | | | |
|---|---|---|---|
| | Registration | In “closed” (non-open) surveys, users need to login first and it is easier to prevent duplicate entries from the same user. Describe how this was done. For example, was the survey never displayed a second time once the user had filled it in, or was the username stored together with the survey results and later eliminated? If the latter, which entries were kept for analysis (eg, the first entry or the most recent)? | See page 6. For the personalised follow-up surveys, a unique survey link was emailed to each participant. Once participants completed their survey, it closed and could not be re-opened again. |
| **Analysis** | Handling of incomplete questionnaires | Were only completed questionnaires analyzed? Were questionnaires which terminated early (where, for example, users did not go through all questionnaire pages) also analyzed? | Two baseline surveys were excluded from the analysis as only the consent question had been answered (see reference to Harrop et al. 2021 on page 7). |
| | Questionnaires submitted with an atypical timestamp | Some investigators may measure the time people needed to fill in a questionnaire and exclude questionnaires that were submitted too soon. Specify the timeframe that was used as a cut-off point, and describe how this point was determined. | No such measures were undertaken. |
| | Statistical correction | Indicate whether any methods such as weighting of items or propensity scores have been used to adjust for the non-representative sample; if so, please describe the methods. | No measures were undertaken to adjust for the non-representative sample. |
