## Supplementary File 3: Template follow-up surveys at T2 and T3 for "Support needs, support use and perceived helpfulness of support in a cohort of people bereaved during the COVID-19 pandemic: Insights from a longitudinal survey"

### Supplementary File 3: Templates for the follow-up surveys at T2 (8-months post-bereavement) and T3 (13-months post-bereavement), respectively, to be personalised in Sections C (types of support used) and D (Ongoing support needs).

**Note:** Where validated measures were used in these surveys (Part A), they are cited rather than reproduced in full. Please refer to the provided reference for details on the instruments' items and scoring instructions.

#### The grief experiences and support needs of people bereaved during Covid-19: Second/Third Survey

Thank you once again for taking part in our study and agreeing to be sent this second/third questionnaire. We really appreciate the time you have taken to help us with this study.

In this second/third questionnaire we are interested to find out more about your grief experiences and wellbeing at this point in time. We would also like to find out about any support that you have been using recently or feel like you may need and any difficulties you may be experiencing accessing support.

More detailed information about the study is included in the document *Information for Participants – Second/Third Survey* that is enclosed in your survey letter. Please read this information to help you decide whether you would like to take part in this second/third survey and complete the consent section below if you would like to continue.

Information about bereavement support services and resources is also provided in the same document and at the end of this survey.

##### Consent

By participating in this survey, you agree that you have read and understood the information provided above and that you are aged 18 or over.

**I confirm that I have read and understood the information provided about the purpose of this study and how my data will be used. I agree to take part in the following survey knowing that all questions are optional and I can finish the survey at any point.**

Please tick the box to agree with the above statement. ☐

#### Thank you for your help!

**Before we start, could you please tell us whether you have experienced any other bereavements of close friends or family members <u>since completing your last questionnaire N months ago</u>?**

| | |
|---|---|
| Yes | ☐ |
| No | ☐ |

**If you have been bereaved again, we are deeply sorry to hear this and appreciate that you may not feel like completing this survey right now. If you would prefer for us to contact you in six months instead, please let us know below and use the enclosed prepaid envelope to return your survey to us.**

| | |
|---|---|
| I would prefer to complete this survey another time. | ☐ |
| I would like to continue with the survey. | ☐ |

| **Comments:** |
|---|

**If you would like to continue, please carry on with the survey.**

#### Part A

This first section contains a series of questions which will help us to understand how your grief is affecting you and how you are coping with and adjusting to your bereavement. Some of these questions you may remember from answering previously, whilst others are new to this questionnaire. For all questions, please remember that there are no right or wrong answers.

**A1. This section of the survey assessed respondents' vulnerability in grief using the validated 9-item Adult Attitude to Grief (AAG) Scale. Details on the AAG and its items can be found here:**

- Sim J, Machin L, Bartlam B. Identifying vulnerability in grief: psychometric properties of the Adult Attitude to Grief Scale. Qual Life Res. 2014 May;23(4):1211-20. doi: 10.1007/s11136-013-0551-1. Epub 2013 Oct 16. PMID: 24129670.

**A2. This section of the survey assessed respondents for symptoms of Prolonged Grief Disorder (PGD) using the 18-item Traumatic Grief Inventory Self-Report version (TGI-SR). Details on the TGI-SR and its items can be found here:**

- Boelen, P. A., & Smid, G. E. (2017). The Traumatic Grief Inventory Self-Report version (TGI-SR): Introduction and preliminary psychometric evaluation. *Journal of Loss and Trauma, 22*(3), 196–212. https://doi.org/10.1080/15325024.2017.1284488
- Boelen PA, Djelantik AAAMJ, de Keijser J, Lenferink LIM, Smid GE. Further validation of the Traumatic Grief Inventory-Self Report (TGI-SR): A measure of persistent complex bereavement disorder and prolonged grief disorder. Death Stud. 2019;43(6):351-364. doi: 10.1080/07481187.2018.1480546. Epub 2018 Jul 17. PMID: 30015568.

**A3. This section of the survey assessed social support using the Inventory of Social Support (ISS), a 5-item measure that aims to capture to what extent a bereaved person can talk with other people about their loss. Details on the ISS items can be found here:**

- Hogan, N. S., & Schmidt, L. A. (2016). Inventory of Social Support (ISS). In R. A. Neimeyer (Ed.), *Techniques of grief therapy: Assessment and intervention* (pp. 99–102). Routledge/Taylor & Francis Group.

**A4. This section of the survey assessed personal wellbeing using the Office for National Statistic (ONS)'s four wellbeing questions (known as ONS4). Details on the ONS4 can be found here:**

- Personal well-being user guidance - Office for National Statistics

#### Part B. Bereavement support

**This section includes questions on your experiences of accessing support and help with your bereavement <u>over the last two months</u>.**

**B1. Over the <u>last two months</u> have you experienced any difficulties getting support for your grief and bereavement?**

| | Yes | Somewhat | No | I've not tried to get their support |
|---|---|---|---|---|
| From friends and family | ☐ | ☐ | ☐ | ☐ |
| From GP surgery | ☐ | ☐ | ☐ | ☐ |
| From bereavement services | ☐ | ☐ | ☐ | ☐ |

**B2. Do any of the responses below describe your experiences <u>over the last two months</u>? (Please select all that apply):**

| | |
|---|---|
| I have not wanted any support from bereavement services because my family and friends provide me with enough support | ☐ |
| I have not wanted any support from bereavement services because I am coping ok without this type of support | ☐ |
| I have not wanted any support from bereavement services because I do not think it would help me | ☐ |
| I do not know how to get support from bereavement services | ☐ |
| I have felt uncomfortable asking for support from bereavement services | ☐ |
| I have felt uncomfortable asking for help or support from friends or family | ☐ |
| The support I wanted from bereavement services was not available to me | ☐ |
| Friends or family have not been able to support me in the way I that wanted | ☐ |

**B3. Please describe any difficulties that you have faced getting support from either friends/family or bereavement services.**

#### Section C. Types of support used

**This section includes questions on the types of support you have been using since becoming bereaved. It includes more detailed questions about support that you may previously have told us about and asks about any new support that you may have been receiving since your last questionnaire.**

**C1. In your previous questionnaire you told us that you have been getting support from [insert place here]. We would like to know a bit more about this support. Firstly, are you still receiving this support?**

| | |
|---|---|
| **Yes**<br>*If yes, please answer questions a-g below.* | ☐ |
| **No**<br>*If no, please tell us when it finished and then answer questions a-g below.*<br>Support no longer received/used from: ………………………………………… | ☐ |

**Can you please also tell us:**

| |
|---|
| a) How is/was the support provided? (e.g. in person, over the phone, via skype/zoom, web-chat etc) |
| b) How effective do (or did) you find this method of communication? |
| c) Roughly when did you first start using this support? |
| d) How often do/did you use this support? If relevant please state the number of sessions received. |
| e) Do you feel that you started using this support at around the right time for you? |

| |
|---|
| f) Do you feel happy with the amount of support you have received from this support provider? |
| g) How did you first find out about this support? |

**You also told us you had been getting support from [insert second source of support from previous survey]. Are you still receiving this support?**

| | |
|---|---|
| **Yes**<br>*If yes, please answer questions a-g below.* | ☐ |
| **No**<br>*If no, please tell us when it finished and then answer questions a-g below.*<br>Support no longer received/used from: ………………………………………… | ☐ |

**Please can you also tell us:**

| |
|---|
| a) How is/was the support provided? (e.g. in person, over the phone, via skype/zoom, web-chat etc) |
| b) How effective do (or did) you find this method of communication? |
| c) Roughly when did you first start using this support? |
| d) How often do/did you use this support? If relevant please state the number of sessions received. |
| e) Do you feel that you started using this support at around the right time for you? |

| |
|---|
| f) Do you feel happy with the amount of support you have received from this support provider? |
| g) How did you first find out about this support? |

**Have you used any other bereavement support services or groups since completing your last survey N months ago?**

| | |
|---|---|
| **Yes**<br>*If yes, please continue to the* ***next question (C2)*** *below.* | ☐ |
| **No**<br>*If no, please continue to* ***Section D on page X.*** | ☐ |

**C2. If you indicated above that you have also used some other support groups, services or resources, please tell us what type of support this is:**

| | |
|---|---|
| Telephone helpline support (e.g. bereavement helpline) | ☐ |
| Online community support via written comments (e.g. Facebook group, online chat forum) | ☐ |
| Informal support/talking group (e.g. social group for bereaved people) | ☐ |
| Formal bereavement support group (e.g. group discussions about bereavement guided by a facilitator; or group counselling) | ☐ |
| One-to-one support (e.g. individual counselling) | ☐ |
| Specialist mental health support | ☐ |
| Other | ☐ |

**If you selected Other, please specify:**

________________________________________________________________________

**Are you still receiving this support?**

| | |
|---|---|
| **Yes**<br>*If yes, please answer questions a-g below.* | ☐ |
| **No**<br>*If no, please tell us when it finished and then answer questions a-g below.*<br><br>Support no longer received/used from: ………………………………………… | ☐ |

**Please can you also tell us the following details:**

| |
|---|
| a) How is/was the support provided? (e.g. in person, over the phone, via skype/zoom, web-chat etc) |
| b) How effective do (or did) you find this method of communication? |
| c) Roughly when did you first start using this support? |
| d) How often do/did you use this support? If relevant please state the number of sessions received. |
| e) Do you feel that you started using this support at around the right time for you? |
| f) Do you feel happy with the amount of support you have received from this support provider? |
| g) How did you first find out about this support? |

**Have you used any other additional bereavement support services, groups or resources since completing your last survey N months ago?**

| | |
|---|---|
| **Yes**<br>*If yes, please continue to the* ***next question (C3)*** *below.* | ☐ |
| **No**<br>*If no, please* ***continue to Section D on page X****.* | ☐ |

**C3. If you indicated above that you have also used some other additional support groups, services or resources, please tell us what type of support this is:**

| | |
|---|---|
| Telephone helpline support (e.g. bereavement helpline) | ☐ |
| Online community support via written comments (e.g. Facebook group, online chat forum) | ☐ |
| Informal support/talking group (e.g. social group for bereaved people) | ☐ |
| Formal bereavement support group (e.g. group discussions about bereavement guided by a facilitator; or group counselling) | ☐ |
| One-to-one support (e.g. individual counselling) | ☐ |
| Specialist mental health support | ☐ |
| Other | ☐ |

**If you selected Other, please specify:**

______________________________________________________________________________

**Are you still receiving/using this support?**

| | |
|---|---|
| **Yes**<br>*If yes, please answer questions a-g below.* | ☐ |
| **No**<br>*If no, please tell us when it finished and then answer questions a-g below.*<br><br>Support no longer received/used from: ………………………………………… | ☐ |

**Please can you also tell us the following details:**

| |
|---|
| a) How is/was the support provided? (e.g. in person, over the phone, via skype/zoom, web-chat etc) |
| b) How effective do (or did) you find this method of communication? |
| c) Roughly when did you first start using this support? |
| d) How often do/did you use this support? If relevant please state the number of sessions received. |
| e) Do you feel that you started using this support at around the right time for you? |
| f) Do you feel happy with the amount of support you have received from this support provider? |
| g) How did you first find out about this support? |

**In addition to the two further sources of support you have just described, are there any other bereavement support services, groups or resources that you have used <u>in the N months since completing your last survey</u>?**

| | |
|---|---|
| **Yes**<br>*If yes, please continue to the* ***next question (C4)*** *below.* | ☐ |
| **No**<br>*If no, please* ***continue to Section D on page X****.* | ☐ |

**C4. If you indicated above that you have also used a third form of additional support, please tell us what type of support this is:**

| | |
|---|---|
| Telephone helpline support (e.g. bereavement helpline) | ☐ |
| Online community support via written comments (e.g. Facebook group, online chat forum) | ☐ |
| Informal support/talking group (e.g. social group for bereaved people) | ☐ |
| Formal bereavement support group (e.g. group discussions about bereavement guided by a facilitator; or group counselling) | ☐ |
| One-to-one support (e.g. individual counselling) | ☐ |
| Specialist mental health support | ☐ |
| Other | ☐ |

**If you selected Other, please specify:**

________________________________________________________________________

**Are you still receiving/using this support?**

| | |
|---|---|
| **Yes**<br>*If yes, please answer questions a-g below.* | ☐ |
| **No**<br>*If no, please tell us when it finished and then answer questions a-g below.*<br><br>Support no longer received/used from: ………………………………………… | ☐ |

**Please can you also tell us the following details:**

| |
|---|
| a) How is/was the support provided? (e.g. in person, over the phone, via skype/zoom, web-chat etc) |
| b) How effective do (or did) you find this method of communication? |
| c) Roughly when did you first start using this support? |
| d) How often do/did you use this support? If relevant please state the number of sessions received. |
| e) Do you feel that you started using this support at around the right time for you? |
| f) Do you feel happy with the amount of support you have received from this support provider? |
| g) How did you first find out about this support? |

#### Section D. Ongoing Support Needs

**This section includes questions on the kinds of support or help that you may have needed over the <u>last two months</u>. It also asks how well you feel these needs have been met by friends/family and any other sources of support you may have been using.**

**D1a. Over the <u>last two months,</u> please tell us how much support or help you have needed with the following:**

| **Practical tasks** | **High level of support needed** | **Fairly high level of support needed** | **Moderate level of support needed** | **Little support needed** | **No support needed** |
|---|---|---|---|---|---|
| Practical and administrative tasks e.g. finances, other paperwork etc. | ☐ | ☐ | ☐ | ☐ | ☐ |
| Getting relevant information and advice e.g. legal, financial, available support | ☐ | ☐ | ☐ | ☐ | ☐ |
| Looking after myself/family e.g. getting food, medication, childcare | ☐ | ☐ | ☐ | ☐ | ☐ |

If you have needed support with the above (*practical tasks*), please tell us how helpful you have found the support of other people, groups or resources in meeting these needs:

| **Sources of support** | **Very helpful** | **Slightly helpful** | **Neither helpful nor unhelpful** | **Slightly unhelpful** | **Very unhelpful** |
|---|---|---|---|---|---|
| Friends and family | ☐ | ☐ | ☐ | ☐ | ☐ |
| [Insert first type of support reported in previous survey] | ☐ | ☐ | ☐ | ☐ | ☐ |
| [Insert second type of support reported in previous survey] | ☐ | ☐ | ☐ | ☐ | ☐ |

| | | | | | |
|---|---|---|---|---|---|
| Other type of support 1<br>*if relevant, please state here:* | ☐ | ☐ | ☐ | ☐ | ☐ |
| Other type of support 2<br>*if relevant, please state here:* | ☐ | ☐ | ☐ | ☐ | ☐ |

Please explain some of the ways you have been helped with your practical support needs. Alternatively, you might like to tell us about any further help that you need with these.

**D2a. Over the <u>last two months,</u> please tell us how much support or help you have needed with the following:**

| **Managing my grief** | **High level of support needed** | **Fairly high level of support needed** | **Moderate level of support needed** | **Little support needed** | **No support needed** |
|---|---|---|---|---|---|
| Dealing with my feelings about being without my loved one | ☐ | ☐ | ☐ | ☐ | ☐ |
| Dealing with my feelings about the way my loved one died | ☐ | ☐ | ☐ | ☐ | ☐ |
| Expressing my feelings and feeling understood by others | ☐ | ☐ | ☐ | ☐ | ☐ |
| Feeling comforted and reassured | ☐ | ☐ | ☐ | ☐ | ☐ |

If you have needed support with the above (managing your grief), please tell us how helpful you have found the support of other people, groups or resources in meeting these needs:

| **Sources of support** | **Very helpful** | **Slightly helpful** | **Neither helpful nor unhelpful** | **Slightly unhelpful** | **Very unhelpful** |
|---|---|---|---|---|---|
| Friends and family | ☐ | ☐ | ☐ | ☐ | ☐ |
| [Insert first type of support reported in previous survey] | ☐ | ☐ | ☐ | ☐ | ☐ |
| [Insert second type of support reported in previous survey] | ☐ | ☐ | ☐ | ☐ | ☐ |
| Other type of support 1<br>*if relevant, please state here:* | ☐ | ☐ | ☐ | ☐ | ☐ |
| Other type of support 2<br>*if relevant, please state here:* | ☐ | ☐ | ☐ | ☐ | ☐ |

Please explain some of the ways in which you have been helped with *managing your grief*. Alternatively, you might like to tell us about any further help that you need with this.

**D3a. Over the last two months, please tell us how much support or help you have needed with the following:**

| **Quality of life and mental wellbeing** | **High level of support needed** | **Fairly high level of support needed** | **Moderate level of support needed** | **Little support needed** | **No support needed** |
|---|---|---|---|---|---|
| Finding balance between grieving and other areas of life | ☐ | ☐ | ☐ | ☐ | ☐ |
| Participating in work, leisure or other regular activities (e.g. shopping, housework) | ☐ | ☐ | ☐ | ☐ | ☐ |
| Managing and maintaining my relationships with friends and family | ☐ | ☐ | ☐ | ☐ | ☐ |
| Loneliness and social isolation | ☐ | ☐ | ☐ | ☐ | ☐ |
| Feelings of anxiety and depression | ☐ | ☐ | ☐ | ☐ | ☐ |
| Regaining sense of purpose and meaning in life | ☐ | ☐ | ☐ | ☐ | ☐ |
| Feeling optimistic and hopeful for the future | ☐ | ☐ | ☐ | ☐ | ☐ |

If you have needed support with the above (quality of life and mental wellbeing), please tell us how helpful you have found the support of other people, groups or resources in meeting these needs:

| **Sources of support** | **Very helpful** | **Slightly helpful** | **Neither helpful nor unhelpful** | **Slightly unhelpful** | **Very unhelpful** |
|---|---|---|---|---|---|
| Friends and family | ☐ | ☐ | ☐ | ☐ | ☐ |
| [Insert first type of support reported in previous survey] | ☐ | ☐ | ☐ | ☐ | ☐ |
| [Insert second type of support reported in previous survey] | ☐ | ☐ | ☐ | ☐ | ☐ |
| Other type of support 1<br>*if relevant, please state here:* | ☐ | ☐ | ☐ | ☐ | ☐ |
| Other type of support 2<br>*if relevant, please state here:* | ☐ | ☐ | ☐ | ☐ | ☐ |

Please explain some of the ways in which you have been helped with your *quality of life and mental wellbeing.* Alternatively, you might like to tell us about any further help that you need with this.

**D4. Do you have any other needs for help or support that have not already been described?**

| | |
|---|---|
| Yes | ☐ |
| No | ☐ |

If yes, please describe:

**D5. Are there any children or young people living with you who have also been affected by this bereavement?**

| | |
|---|---|
| Yes | ☐ |
| No | ☐ |

If yes, how old are they?

……………………………………………………………………………………………………………………………………………

**Please tell us about any support that you feel they need and/or any support they have been receiving:**

**D6. Do you feel that there are any changes to government policy or support services that could be made to help you at this time (and which you have not already described)?**

**D7. If you would like to write anything further about your experiences of grieving over the last few months and/or how you feel you have been coping please use the box below:**

#### Section E.

**In this section we would like to ask you a few additional questions about other aspects of your life that may affect or have been affected by your bereavement.**

**E1. Have you been diagnosed with any illness or medical condition since you completed the previous survey N months ago?**

| | |
|---|---|
| Yes | ☐ |
| No | ☐ |

If yes, please provide details:

……………………………………………………………………………………………………………………………………………

**E2. Roughly how many appointments with your GP have you had over the last two months?**

……………………………………………………………………………………………………………………………………………

**E3. Roughly how many times have you bought over-the-counter medicine (e.g. paracetamol, sleep aids or stress relief remedies) over the last two months?**

| | |
|---|---|
| Never | ☐ |
| One to three times | ☐ |
| 4 times or more | ☐ |

**E4. Have you experienced any difficulties with sleeping over the last two months?**

| | |
|---|---|
| Yes | ☐ |
| No<br>*If no, please continue to question E5.* | ☐ |

**If yes, how often do you have difficulties with sleeping (e.g. trouble falling asleep, waking up early or in the middle of the night, bad dreams, poor overall sleep quality)?**

| | |
|---|---|
| Less than once a week | ☐ |
| Once or twice a week | ☐ |

| | |
|---|---|
| Three or more times a week | ☐ |

**To enable us to better understand how bereavement impacts upon the employment and working life of bereaved people, please answer the following questions.**

**E5. Has your employment status changed since your last survey N months ago?**

| | |
|---|---|
| **Yes** | ☐ |
| **No** | ☐ |
| **Not relevant for me**<br>*(e.g. retired, full-time student, permanently sick/disabled, long-term unemployed, looking after the home, caring for a loved one)*<br>**If not relevant for you, please continue to the end of the survey on page XX.** | ☐ |

If yes, please provide details (including if you have been furloughed):

……………………………………………………………………………………………………………………………

**E6. Approximately how much time have you had off work since becoming bereaved (in days/weeks or months)?**

| |
|---|
| Amount of time off work due to bereavement, illness or stress: |
| Amount of time off work due to furlough or unemployment: |

| |
|---|
| Amount of time off work due to other extended periods of leave (please specify below): |

If you indicated above that you had time off work for other reasons (e.g. school holidays/closures, caring responsibilities, maternity leave), please specify here:

……………………………………………………………………………………………………………………………………………

**E7. If you would like to share further information about your time off work since your bereavement, please use the box below:**

**Many thanks for completing this second survey and for all of the time that you have given to this research so far.** Your responses will help to enable others who are bereaved to access the support they need. We appreciate that this may have been difficult and painful for you and we are very grateful for your contribution.

We would like to send you one final questionnaire six/twelve months from now, which will enable us to consider the longer-term impacts of bereavement during the Covid-19 pandemic. Please use the box below to tell us if you are happy for us to send you one final questionnaire.

| | |
|---|---|
| Yes | ☐ |
| No | ☐ |

Thank you again for your help. We are extremely grateful for your contribution.

Here are our contact details should you wish to get in touch: [name, email address and phone number]

If you would like to talk to somebody about your bereavement, you can access support from these services:

- Marie Curie Bereavement Support: 0800 090 2309 https://www.mariecurie.org.uk/help/support/bereaved-family-friends/dealing-grief/bereavement-or-grief-counselling
- Cruse Bereavement Care: 0808 808 1677 https://www.cruse.org.uk/
- NHS Bereavement Helpline: 0800 2600 400 https://www.nhs.uk/conditions/stress-anxiety-depression/coping-with-bereavement/
- The Good Grief Trust: https://www.thegoodgrieftrust.org/
- At a Loss: www.ataloss.org
- The National Bereavement Service (NBS): https://www.thenbs.org/

#### *Thank you again for your help!*
