## Supplementary File 4: Template follow-up survey at T4 for "Support needs, support use and perceived helpfulness of support in a cohort of people bereaved during the COVID-19 pandemic: Insights from a longitudinal survey"

### Supplementary File 4: Follow-up survey at T4 (25-months post-bereavement)

**Note:** Where validated measures were used in this survey (Part A), they are cited rather than reproduced in full. Please refer to the provided reference for details on the instruments' items and scoring instructions.

#### The grief experiences and support needs of people bereaved during Covid-19: Final Survey

Thank you once again for taking part in our study and agreeing to be sent this final questionnaire. We really appreciate the time you have taken over the last 18 months to help us with this study. Information from this final survey will be a great help in understanding longer-term experiences of grief and bereavement at this challenging time.

In this questionnaire we are interested to find out more about your grief experiences and wellbeing at the present time. We would also like to find out:

- What support you have been using recently (including informal support from family and friends).
- Any difficulties you may have experienced accessing support.
- What other type of support you feel you may still need.

More detailed information about the study is included in the document *Information for Participants – Final Survey* that is enclosed with your survey invitation. Please read this information to help you decide whether you would like to take part in this final survey and complete the consent section below if you would like to continue.

Information about bereavement support services and resources is also provided in the same document and at the end of this survey.

##### Consent

By participating in this survey, you agree that you have read and understood the information provided above and that you are aged 18 or over.

**I confirm that I have read and understood the information provided about the purpose of this study and how my data will be used, including the potential publication of my anonymised quotations to illustrate research findings. I agree to take part in the following survey knowing that all questions are optional and I can finish the survey at any.**

- Personal well-being user guidance - Office for National Statistics

**A5. If you would like to tell us a bit more about how you are currently coping with regards to the loss of your [insert], please use the box below.**

#### Part B. Bereavement support

**This section includes questions on your support needs and experiences of accessing support and help with your bereavement.**

**B1. Over the last TWO MONTHS have you needed support with the following?:**

| | High level of support needed | Fairly high level of support needed | Moderate level of support needed | Little support needed | No support needed |
|---|---|---|---|---|---|
| Practical tasks relating to the death e.g. sorting out affairs, paperwork etc. | ☐ | ☐ | ☐ | ☐ | ☐ |
| Getting relevant information and advice e.g. legal, financial, available support | ☐ | ☐ | ☐ | ☐ | ☐ |
| Looking after myself/family e.g. getting food, medication, childcare etc | ☐ | ☐ | ☐ | ☐ | ☐ |
| Dealing with my feelings about being without my loved one | ☐ | ☐ | ☐ | ☐ | ☐ |
| Dealing with my feelings about the way my loved one died | ☐ | ☐ | ☐ | ☐ | ☐ |
| Expressing my feelings and feeling understood by others | ☐ | ☐ | ☐ | ☐ | ☐ |
| Feeling comforted and reassured | ☐ | ☐ | ☐ | ☐ | ☐ |
| Loneliness and social isolation | ☐ | ☐ | ☐ | ☐ | ☐ |
| Managing and maintaining my relationships with friends and family | ☐ | ☐ | ☐ | ☐ | ☐ |
| Finding balance between grieving and other areas of life | ☐ | ☐ | ☐ | ☐ | ☐ |
| Participating in work, leisure or other regular activities (e.g. shopping, housework) | ☐ | ☐ | ☐ | ☐ | ☐ |
| Feelings of anxiety and depression | ☐ | ☐ | ☐ | ☐ | ☐ |

| | High level of support needed | Fairly high level of support needed | Moderate level of support needed | Little support needed | No support needed |
|---|---|---|---|---|---|
| Regaining sense of purpose and meaning in life | ☐ | ☐ | ☐ | ☐ | ☐ |

**Please provide further details on these or any other areas of support that you feel you have needed over the last six months:**

**B2. Over the last six months have you experienced any difficulties getting support for your grief and bereavement?**

| | Yes | Somewhat | No | I've not tried to get their support |
|---|---|---|---|---|
| From friends and family | ☐ | ☐ | ☐ | ☐ |
| From GP surgery | ☐ | ☐ | ☐ | ☐ |
| From bereavement services | ☐ | ☐ | ☐ | ☐ |

**B3. Do any of the responses below describe your experiences <u>over the last six months</u>? (Please select all that apply):**

| | |
|---|---|
| I have not wanted any support from bereavement services because my family and friends provide me with enough support | ☐ |
| I have not wanted any support from bereavement services because I am coping ok without this type of support | ☐ |
| I have not wanted any support from bereavement services because I do not think it would help me | ☐ |
| I feel that the support that is available from bereavement services is not appropriate for my needs | ☐ |
| I do not know how to get support from bereavement services | ☐ |
| I have felt uncomfortable asking for support from bereavement services | ☐ |
| I have felt uncomfortable asking for help or support from friends or family | ☐ |
| The support I wanted from bereavement services was not available to me | ☐ |
| Friends or family have not been able to support me in the way I that wanted | ☐ |

**B4. Please tell us more about these or any other difficulties that you have faced getting support from either friends/family or bereavement services <u>in the last six months</u>.**

**B5. Are there any children or young people under the age of 25 living with you who have also been affected by this bereavement?**

| | |
|---|---|
| Yes | ☐ |
| No | ☐ |

**a. How old are they?**

……………………………………………………………………………………………………………………………………………

**b. Please tell us about any support that you feel they need and/or any support they have been receiving <u>during the last six months</u>:**

**B6. Apart from any children or young people who might live with you, are there any other people close to you who have been particularly affected by this bereavement?**

#### Section C. Types of support used

**This section includes questions on the types of support you have been using over the last SIX months.**

**C1. What types of support or resources have you used over the last six months to help you cope with your bereavement**? (Please select all that apply)

| | |
|---|---|
| **Friend or family** | ☐ |
| **Written or audio resources** (e.g. self-help guides, books or websites, podcasts) | ☐ |
| **GP or other member of staff at the GP surgery** | ☐ |
| **Telephone helpline support** (e.g. bereavement helpline) | ☐ |
| **Instant webchat support service** (e.g. exchanging written webchat messages with a trained or professional support provider in real time) | ☐ |
| **Online bereavement community support via written comments** (e.g. Facebook group, online chat forums with other bereaved people) | ☐ |
| **Community groups** (groups with a focus on social/recreational activities rather than bereavement, e.g. faith groups, reading groups, gardening groups) | ☐ |
| **Informal bereavement support group** (e.g. peer support group for bereaved people) | ☐ |
| **Formal bereavement support group** (e.g. group discussions about bereavement guided by a trained or professional facilitator; or group counselling) | ☐ |
| **One-to-one support** (e.g. individual counselling) | ☐ |
| **Specialist mental health support** | ☐ |

**If you selected "Community groups" above, how was this support provided?**

| | |
|---|---|
| In person | ☐ |
| Virtually via group calls (via e.g. Zoom, Skype etc.) | ☐ |
| A mix of both | ☐ |

**If you selected "Informal bereavement support group" above, how was this support provided?**

| | |
|---|---|
| In person | ☐ |
| Virtually via group calls (via e.g. Zoom, Skype etc.) | ☐ |
| A mix of both | ☐ |

**If you selected "Formal bereavement support group" above, how was this support provided?**

| | |
|---|---|
| In person | ☐ |
| Virtually via group calls (via e.g. Zoom, Skype etc.) | ☐ |
| A mix of both | ☐ |

**If you selected "One-to-one support" above, how was this support provided?**

| | |
|---|---|
| In person | ☐ |
| Virtually via group calls (via e.g. Zoom, Skype etc.) | ☐ |
| A mix of both | ☐ |

**If you selected "Specialist mental health support" above, how was this support provided?**

| | |
|---|---|
| In person | ☐ |
| Virtually via group calls (via e.g. Zoom, Skype etc.) | ☐ |
| A mix of both | ☐ |

**C2. Please tell us which of these type(s) of support you have found most helpful <u>in the last six months</u> and how the support has helped you.**

**C3. Have you used any other types of support to those listed above?**

| | |
|---|---|
| Yes | ☐ |
| No | ☐ |

**If yes, please tell us what support this is and how it has helped you.**

**To help plan support services going forwards we would like to find out about your preferences for the different types of bereavement support that are available.**

**C4. Please tell us how much you would like or appreciate each of the following types of support if you were to experience another close bereavement in non-pandemic circumstances (i.e. when we can meet freely with others because infection control measures are no longer needed).**

1. **Self-help resources and informal social support**

| | Strongly like | Quite like | Neither like nor dislike | Slightly dislike | Strongly dislike |
|---|---|---|---|---|---|
| **Friends or family** | ☐ | ☐ | ☐ | ☐ | ☐ |
| **Written or audio resources** (e.g. self-help guides, books or websites, podcasts) | ☐ | ☐ | ☐ | ☐ | ☐ |
| **Online bereavement community support via written comments** (e.g. Facebook group, online chat forums with other bereaved people) | ☐ | ☐ | ☐ | ☐ | ☐ |
| **Community groups** (e.g. groups with a focus on social/recreational activities rather than bereavement e.g. faith groups or reading groups, gardening groups) **– IN PERSON MEETINGS** | ☐ | ☐ | ☐ | ☐ | ☐ |
| **Community groups** (e.g. groups with a focus on social/recreational activities rather than bereavement e.g. faith groups or reading groups, gardening groups) **– VIRTUAL MEETINGS VIA VIDEO CALL** | ☐ | ☐ | ☐ | ☐ | ☐ |
| **Informal bereavement support group** (e.g. peer support group for bereaved people) **- IN PERSON MEETINGS** | ☐ | ☐ | ☐ | ☐ | ☐ |
| **Informal bereavement support group** (e.g. peer support group for bereaved people) **– VIRTUAL MEETINGS VIA VIDEO CALLS** | ☐ | ☐ | ☐ | ☐ | ☐ |

**2. Support from GPs and helplines/instant webchat services**

| | Strongly like | Quite like | Neither like nor dislike | Slightly dislike | Strongly dislike |
|---|---|---|---|---|---|
| **Bereavement conversation with GP or other member of practice staff** (including signposting and/or timely referrals to support) | ☐ | ☐ | ☐ | ☐ | ☐ |
| **Telephone helpline support** (e.g. bereavement helpline) | ☐ | ☐ | ☐ | ☐ | ☐ |
| **Instant webchat support service** (e.g. exchanging written webchat messages with a trained or professional support provider in real time) | ☐ | ☐ | ☐ | ☐ | ☐ |

**3. Formal Bereavement and Mental Health Support**

| | Strongly like | Quite like | Neither like nor dislike | Slightly dislike | Strongly dislike |
|---|---|---|---|---|---|
| **Formal bereavement support group** (e.g. group counselling/discussions about bereavement guided by a trained or professional facilitator) **– IN MEETINGS** | ☐ | ☐ | ☐ | ☐ | ☐ |
| **Formal bereavement support group** (e.g. group counselling/discussions about bereavement guided by a trained or professional facilitator) **– VIRTUAL MEETINGS VIA VIDEO CALL** | ☐ | ☐ | ☐ | ☐ | ☐ |
| **One-to-one support** (e.g. individual grief counselling) – **PROVIDED IN PERSON** | ☐ | ☐ | ☐ | ☐ | ☐ |
| **One-to-one support** (e.g. individual grief counselling) – **PROVIDED VIA TELEPHONE OR VIDEO CALL** | ☐ | ☐ | ☐ | ☐ | ☐ |
| **Specialist mental health support** – **PROVIDED IN PERSON** | ☐ | ☐ | ☐ | ☐ | ☐ |

| | | | | | |
|---|---|---|---|---|---|
| **Specialist mental health support – PROVIDED VIA TELEPHONE OR VIDEO CALL** | ☐ | ☐ | ☐ | ☐ | ☐ |

**C5. Please use the box below to tell us more about your preferences for these different types of support or any other type of support not on our list that you think would be helpful.**

**C6. Thinking back on the time since [insert] died, what aspect(s) of your bereavement or grief have you found the most challenging?**

**C7. What helped or would have helped you the most, to cope and adjust over this period of time?**

#### Section D.

#### In this section we would like to ask you a few additional questions about other aspects of your life that may affect or have been affected by your bereavement.

**D1. Have you been diagnosed with any illness or medical condition <u>in the past year</u>?**

| | |
|---|---|
| Yes | ☐ |
| No | ☐ |

If yes, please provide details:

……………………………………………………………………………………………………………………………………………

**D2. Roughly how many appointments with your GP have you had over <u>the last two months</u>?**

……………………………………………………………………………………………………………………………………………….

**D3. Roughly how many times have you bought over-the-counter medicine (e.g. paracetamol, sleep aids or stress relief remedies) over the <u>last two months</u>?**

| | |
|---|---|
| Never | ☐ |
| One to three times | ☐ |
| 4 times or more | ☐ |

**D4. Have you experienced any difficulties with sleeping over the <u>last two months</u>?**

| | |
|---|---|
| Yes | ☐ |
| No<br>*If no, please continue to question E5.* | ☐ |

**If yes, how often do you have difficulties with sleeping (e.g. trouble falling asleep, waking up early or in the middle of the night, bad dreams, poor overall sleep quality)?**

| | |
|---|---|
| Less than once a week | ☐ |
| Once or twice a week | ☐ |
| Three or more times a week | ☐ |

**To enable us to better understand how bereavement impacts upon the employment and working life of bereaved people, please answer the following questions.**

**D5. Has your employment status changed <u>in the last 12 months</u>?**

| | |
|---|---|
| **Yes** | ☐ |
| **No** | ☐ |
| **Not relevant for me**<br>*(e.g. retired, full-time student, permanently sick/disabled, long-term unemployed, looking after the home, caring for a loved one)*<br>**If not relevant for you, please continue to the end of the survey on page 21.** | ☐ |

If yes, please briefly describe in what way your employment status has changed:

**Is the change in your employment status in any way related to your bereavement (e.g. you have reduced your hours or changed jobs)?**

| | |
|---|---|
| Yes | ☐ |
| No | ☐ |

**D6. Have you had any time off work <u>during the last twelve months</u> due to bereavement, illness or stress?**

| | |
|---|---|
| **Yes** | ☐ |
| **No** | |
| **Not relevant for me**<br>*(e.g. retired, full-time student, permanently sick/disabled, long-term unemployed, looking after the home, caring for a loved one)* | ☐ |

**If yes, approximately how much time have you had off work due to bereavement, illness or stress (in days, weeks or months?)**

| |
|---|
| Amount of time off work due to bereavement, illness or stress: |

**D7. If you would like to share further information about your time off work <u>in the last twelve months</u>, please use the box below:**

**Many thanks for completing this final survey and for all of the time that you have given to this research.** Your responses will help to enable others who are bereaved to access the support they need. We appreciate that this may have been difficult and painful for you and we are very grateful for your contribution.

Before you submit your completed survey, please indicate below if you would like to receive a final summary of the results and an end-of-study update.

**I would like to receive a final study update and result summary.**

| | |
|---|---|
| Yes | ☐ |
| No | ☐ |

Should you wish to get in touch, please find our contact details here [contact details].

If you would like to talk to somebody about your bereavement, you can access support from these services:

- Marie Curie Bereavement Support: 0800 090 2309 https://www.mariecurie.org.uk/help/support/bereaved-family-friends/dealing-grief/bereavement-or-grief-counselling
- Cruse Bereavement Care: 0808 808 1677 https://www.cruse.org.uk/
- NHS Bereavement Helpline: 0800 2600 400 https://www.nhs.uk/conditions/stress-anxiety-depression/coping-with-bereavement/
- The Good Grief Trust: https://www.thegoodgrieftrust.org/
- At a Loss: www.ataloss.org

### Thank you again for your help.
