## Supplementary File 6: Full themes table for "Support needs, support use and perceived helpfulness of support in a cohort of people bereaved during the COVID-19 pandemic: Insights from a longitudinal survey"

**Supplementary File 6:** Overview of qualitative themes (mapped onto the three support domains 'Practical support', 'Managing grief' and 'Quality of life/mental wellbeing') with illustrative quotes from across the four survey time points and different support types.

| | Main themes | Sub-themes | Illustrative quotes |
|---|---|---|---|
| **Practical support** | Help with looking after oneself, family and home | • Tangible help with chores<br>• Help with looking after dependents | "I have felt overwhelmed with daily tasks such as shopping and housework and family have been supporting me with this." [Family and friends] (PID 053 at T2)<br><br>"Partner and one of my daughters provide an ear to listen when I'm weepy and help with some of the day to day running of the house when I doubt feel up to it. [Family and friends] (PID 541 at T3) |
| | Help with bereavement-related practical and administrative tasks | • Organising funerals, memorials etc.<br>• Dealing with probate and other paperwork<br>• Dealing with deceased's possessions and selling properties<br>• Advice on legal, financial or administrative questions<br>• Advice on how/where to get support | "Daughter and son in law sorted all the paperwork out for me and helped with the funeral arrangements and notifying family and friends." [Family and friends] (PID 224 at T2)<br><br>"My partner does a lot of cooking for me and helps me with finances and when we had to sell my parent's house. I also had some friends help me clear my parent's house." [Family and friends] (PID 323 at T2)<br><br>"Facebook group has provided helpful links and resources." [Online community support] (PID 094 at T1) |
| **Managing grief** | Expressing feelings and feeling understood | • Being able to talk about feelings and feel listened to, understood and accepted<br>• Sharing with others with similar experiences<br>• Benefits of sharing with independent strangers | "Just by having a partner that would listen has been amazing. Grief creeps up unexpectedly and having someone who is prepared to listen and with whom I can express my sorrow helps." [Family and friends] (PID 325 at T2)<br><br>"Being allowed to talk to friends with shared experiences, without anyone thinking "she should be over it by now" "oh no I can't hear all this again" sharing with people who understand. Friends and family can only do so much as they have never experienced child loss." [Family and friends] (PID 273 at T2)<br><br>"People listening, especially when I get upset." [Family and friends] (PID 351 at T3)<br><br>"Family that aren't afraid to discuss our grief. Friends who also made an effort to talk about grief." [Family and friends] (PID 394 at T4)<br>"People who have lost people too, so they understand the grief. Easier to vent and share feelings |

| | | | |
|---|---|---|---|
| | | | there." [Online community support] (PID 089, at T2)<br><br>"The groups on Facebook because they had suffered the same kind of loss they understand and they just listen and send advice on how they managed and sometimes is just the hope they give... The psychologist is always there to listen and to help me to work my grief. I don't need to look OK or strong to her or to the Facebook group people. But I don't want to worry my friends and family so I hide how I am most of the times." [Online community support/One-to-one support] (PID 088 at T3)<br><br>"They were great, they just listened and let me cry / vent, that is what I needed." [Helplines & instant webchat services] (PID 513 at T2)<br><br>"I feel the counselling has helped, being able to talk to someone who doesn't know you has been beneficial, no judgement. Friends and family too listen but I don't always feel I can go into great detail with them as it maybe upsetting for them. So I tend not to do that, maybe I don't want them to feel the pain I feel. But with the counsellor, she has no connection to my son. So I can tell her things I maybe couldn't tell my family." [One-to-one support] (PID 275 at T4)<br><br>"The widows club is useful as you can say how you feel and know that they understand as they have been through the same thing." [Informal peer-support group] (PID 594 at T2)<br><br>"Connecting with others in a similar situation very helpful-not having to feel like you have to 'explain yourself' or validate your feelings." [Formal bereavement support group] (PID 179 at T2) |
| | Understanding and accepting grief and loss | • Learning about and understanding the grieving process<br>• Normalising and accepting feelings<br>• Recognising that feelings are normal and not alone feeling this way | "Knowing what/how I'm feeling is normal. That others feel the same." [Online community support] (PID 220 at T2)<br><br>"Having the one-to-one counselling has really helped with managing my grief. It has helped me to understand the complexity, duration and feelings I am having. It has been a space to think out loud, to reflect and express myself." [One-to-one support] (PID 563 at T2)<br><br>"Grief counselling has helped me express my feelings without putting a burden on family. Have been able to explore my feelings, be reassured they are normal, and find ways to further process and come to terms with events." [One-to-one support] (PID 115 at T3)<br><br>"I found articles on how men and women grieve differently to be useful as my way of grieving |

| | | | |
|---|---|---|---|
| | | | was quite different from my husband's and it caused some extra stress and pain for several weeks during the early days." [Self-help resources] (PID 429 at T1)<br><br>"It [podcast] has been something I can listen to while out walking and hear other people's experiences. Helps me to process my own feelings and recognise that what I'm feeling is normal." [Self-help resources] (PID 070 at T2) |
| | Remembering and maintaining a connection to the person who has died | • Talking about the person who has died, reminiscing, sharing positive memories<br>• Activities to maintain connection to the person who has died and keep memory alive (includes self-initiated activities) | "My children have allowed me to talk about my Mum (their Nana) and we are now able to start remembering the happy times and not just the trauma of her passing." [Family and friends] (PID 178 at T3)<br><br>"Talking about my son to others, and sharing memories, both of his illness and his life before he became so unwell helps get the balance right." [Family and friends] (PID 527 at T3)<br><br>"The [name of group] fb page - I know it helps me keep his memory alive." [Online community support] (PID 251 at T3)<br><br>"It's helpful to have access to a group of people at work who understand my situation and who I can have a virtual cup of tea with, a good cry or a good laugh as we share memories." [Informal peer-support group] (PID 694 at T1) |
| **Quality of life and mental wellbeing** | Feeling connected to a caring and supportive social network | • Feeling connected to and comforted by a caring and kind others<br>• Feeling seen in one's grief and having experience acknowledged: Knowing that others care.<br>• Mutuality of support | "I have found friends and family just being there and knowing there are people I could call at any time of day or night if I needed to has been probably the best help." [Family and friends] (PID 284 at T1)<br><br>"Friends coming to me of their own accord and making sure I know they care has helped too." [Family and friends] (PID 057 at T2)<br><br>"It has saved me to know that I am not alone in the horror of this grief experience. People understand and care." [Online community support] (PID 251 at T2)<br><br>"I use the social media sites as I feel that the people on the sites have gone through the same as myself and understand my feelings, none of them are judgemental and most of the time I get lovely support messages from individuals, no one on the sites have said it has been over a year so get over it, they have all been very lovely and I hope that my messages to others help them too." [Online community support] (PID 232 at T3) |

| | | | |
|---|---|---|---|
| | | | "I feel that I have made further friends through the zoom and independent meetings that we as a group have started, and as a result my fellow bereaved associates have really helped. Particularly at weekends or in very dark moments, as we are in the same situation, and don't have to apologise when we get really emotional. There is a deep understanding and nonjudgemental attitude. We literally laugh, cry and support each other through companionship, trust and compassion." [Informal peer-support group] (PID 709 at T3) |
| | Managing and finding meaning in life | • Company and inclusion in activities<br>• Distraction: Focus on something other than grief<br>• Encouragement and support for self-care activities<br>• Reassurance for feelings of anxiety and self-doubt<br>• Supportive and understanding work environments<br>• Looking forward and finding hope.<br>• Help with decision-making<br>• Addressing questions of meaning and purpose | "Friends make me do things, get out, join in and although sometimes I don't want to...I usually feel better when I have joined in." [Family and friends] (PID 081 at T2)<br><br>"Chatting generally about the future with friends and making plans for the future." [Family and friends] (PID 498 at T3)<br><br>"I am lucky enough I have a couple of fantastic male platonic friends who have really helped. I went back to meditation and one encourages me with that. The other goes to the gym with me and long walks." [Family and friends] (PID 380 at T4)<br><br>"I found for that reason meeting up with other widows has been good because you can say how you feel and people understand. Not that is all we talk about and go to the theatre and lunch." [Informal peer-support group] (PID 594 at T4)<br><br>"The groups on Facebook I can just vent all what I feel there and I always have a nice word from someone there. There is always someone to share experiences and to give you hope." [Online community support] (PID 088 at T2)<br><br>"My counsellor helped me make decisions about practical things like selling the house." [One-to-one support] (PID 323 at T2)<br><br>"I was quite low in mood and required counselling to allow me to function and regain some control of my mood and life purpose initially it put a strain on my marriage but we have worked through the difficulty." [One-to-one support] (PID 083 at T1)<br><br>"Work have been incredibly supportive. Did a 6-week passed return and although back to usual hours I am taking a day's annual leave a week until Christmas. Have regular 1:1 with my manager to check I am OK." [Workplaces] (PID 081 at T2) |
| | Managing mental | • Self-care activities to improve wellbeing. | "Talking with a therapist has been very helpful. I had a period of depression that triggered my seeking professional help due to suicidal thoughts and feelings. This has enabled me to better |

| | | | |
|---|---|---|---|
| | wellbeing and mental health needs | • Managing self-expectations: Accepting attitudes towards feelings and giving oneself time and space.<br>• Help with managing other challenging life events whilst grieving.<br>• Help for coping with anxiety, stress, depression/low mood, trauma and suicidal thoughts | manage my emotions and to find some meaning in life after [name]'s death." [One-to-one support] (PID 122 at T2)<br><br>"My therapist has helped me to manage the enormous number of adverse situations that have cropped up at once over the last year or two. […] My most significant current difficulties are not directly related to grief, but the loss of my dad has certainly had an impact on the way that I receive, process and cope with other bad news." [One-to-one support] (PID 141 at T2)<br><br>"Counselling suggested strategies to try, but perhaps most of all encouraged me just to accept that feeling down and upset is ok and to be kinder to myself." [One-to-one support] (PID 351 at T3) |
